# Trends in hospitalisations for lower respiratory tract infection after the COVID-19 pandemic in adults with chronic respiratory disease

**DOI:** 10.1101/2024.07.23.24310871

**Authors:** Alexandre Sabaté-Elabbadi, Lucie Brolon, Christian Brun-Buisson, Didier Guillemot, Muriel Fartoukh, Laurence Watier

## Abstract

**Introduction:** COVID-19 pandemic has modified the epidemiology of lower respiratory tract infections (LRTI), particularly in patients presenting a chronic respiratory disease (CRD). LRTI incidence substantially decreased at the start of the COVID-19 pandemic. However, studies focusing on the post-pandemic period are missing. We aimed to evaluate the impact of the pandemic and post-pandemic periods on hospital admissions for LRTI, with a focus on patients with CRD.

**Methods:** From July 2013 to June 2023, monthly numbers of adult hospitalisations for LRTI (excluding SARS-CoV-2) were extracted from the anonymized French National Hospital Discharge Database. They were modelled by regressions with autocorrelated errors. Three periods were defined: (1) early pandemic and successive lockdowns (April 2020 to May 2021); (2) gradual lifting of restrictions and widespread SARS-CoV-2 vaccination (June 2021 to June 2022); (3) withdrawal of restriction measures (July 2022 to July 2023). Analyses were computed for the entire series, by gender, age, severity, and pre-existing CRD

**Results:** Before the pandemic, LRTI hospitalisations showed a winter seasonal pattern with a rising trend. Pre-pandemic incidence was 96 (90.5 to 101.5) per 100,000 population. Compared with the pre-pandemic period, seasonality was no longer present and significant reductions were estimated in the first two periods: −43.64% (−50.11 to −37.17) and −32.97% (−39.88 to −26.05), respectively. A rebound with a positive trend and a seasonal pattern was observed in period 3. Similar results were observed for CRD patients with no significant difference with pre-pandemic levels in the last period (−9.21%; −20.9% to 1.67%), albeit with differential changes according to the type of CRD.

**Conclusions:** COVID-19 pandemic containment measures contributed to significant changes in LRTI incidence, with a rapid increase and return to a seasonal pattern after their gradual lifting, particularly in patients with CRD.

## INTRODUCTION

Community-acquired lower respiratory tract infections (LRTI) encompass heterogeneous conditions of community onset, including community-acquired pneumonia (CAP), and other respiratory tract infections (acute bronchitis, acute exacerbation of chronic obstructive pulmonary disease (COPD), acute exacerbation of bronchiectasis) (1). LRTI remain the major cause of death from infectious diseases worldwide (2). In Europe, the burden is substantial with an incidence of 1.5 per 1,000 inhabitants, and the economic impact is considerable, with an estimated annual cost in Europe of 10 billion Euros (3). In addition, patients with chronic respiratory disease (CRD) are particularly at risk of respiratory infection (4,5), and related mortality (6).

The public health measures implemented in many countries to contain COVID-19 pandemic led to a substantial drop in hospitalisations for other causes of LRTI (7,8). This decline was partly driven by a decrease in the circulation of seasonal respiratory viruses (8,9), but also by a concomitant reduction in the number of bacterial infections associated with microorganisms usually involved in LRTI (10). This reduction in hospital admissions for LRTI was also observed in patients with CRD (11,12), as respiratory viruses are known to contribute significantly to CRD exacerbations in this population (13).

In France, as in many European countries, non-pharmaceutical interventions (NPI) varied during the pandemic (14); with particularly strict measures in 2020 until the spring of 2021, then a gradual relaxation of these measures until August 2022 when they were totally withdrawn (15). Data suggest that gradual easing of NPI has been accompanied by a resurgence in respiratory-transmitted viruses (16), notably respiratory syncytial virus (RSV) (17). However, studies analysing the overall changes in hospitalisations for LRTI associated with these NPI and their lifting during and after the pandemic, are lacking, particularly in patients with CRD. Moreover, if COVID-19 is evolving into a seasonal pattern as research suggests (18), it is crucial to gather data on its post-pandemic impact on the overall LRTI burden.

We aimed to assess and quantify the impact of the pandemic on the trend of hospitalisations for LRTI and the characteristics and severity of inpatients, both overall and among those with a preexisting CRD, during and after the pandemic.

## METHODS

### Study population and selection of hospital stays with LRTI

Hospitalisations of patients aged > 15 years with LRTI (excluding SARS-CoV-2) in metropolitan France from July 2013 to June 2023 were extracted and aggregated monthly, from the French National Hospital Discharge Database (PMSI: Programme de Médicalisation des Sytèmes d’Informations) (19). LRTI was defined by the presence of a related ICD-10 code during hospital stay. Rates of hospitalisations per 100,000 inhabitants and per month were calculated, taking into account the French population growth. For each stay, gender, age, type (acute pneumonia, acute bronchitis, acute exacerbation of COPD, acute exacerbation of bronchiectasis) and severity of LRTI (moderate, severe, and death) (20), were also extracted. Presence of a CRD was defined by an ICD-10 code during the hospital stay. For more details on population selection, see **Table S1-3, Supplementary data**. We then classified the CRD into 8 non-exclusive categories representing the main CRD: (i) COPD, (ii) Asthma, (iii) Cystic Fibrosis and other bronchiectasis (CF-Bronchiectasis), (iv) interstitial lung disease (ILD), (v) thoracic oncology, (vi) pulmonary vascular disease (PVD), (vii) CRD associated with neuromuscular disease (CRD-NM), and (viii) lung transplantation.

Finally, we also considered monthly numbers of hospitalisations with LRTI including SARS-CoV-2 in the last period (i.e. without restriction measures).

### Ethics Statement

As this was a retrospective study of an anonymized database, ethics committee approval was not required, as per French law. The study was conducted using the Institut National de la Santé et de la Recherche Médicale permanent access to the the French National Hospital Discharge Database (PMSI).

### Statistical analyses

To study the impact of the pandemic periods on changes in hospitalisations rates for LRTI, segmented regression models with autocorrelated errors were used (21). Three periods were defined: the first covers the start of the pandemic in France, including 3 successive lockdowns (April 2020 to May 2021); the second covers the gradual removal of restrictions and the widespread COVID-19 vaccination (June 2021 to June 2022); and the last, extending from July 2022 to July 2023, covers the last epidemic season without restriction measures **(Table S4, Supplementary data)**. For each period, a regression line with a trigonometric function, if the series exhibited annual seasonality, was estimated. If non-significant, the slope was removed. We quantified the estimated difference in incidence and percent changes of monthly rates of hospitalisations for each intervention, as compared with the pre-pandemic period, by calculating the difference between the monthly estimated incidence predicted by the model and the one expected under the assumption of no change since the pre-pandemic period. More details are available in **Supplementary data**.

Analyses were performed for LRTI excluding SARS-CoV-2, by age group (15 to 54, 55 to 64, 65 to 74 and >75 years), gender, type of LRTI (classified as pneumonia or other types of LRTI), severity, and presence of a CRD. A final analysis was related to the LRTI series including SARS-CoV-2 in the last period.

Statistical analyses were computed with SAS^®^ Enterprise Guide (Version 8.3; SAS Institute Inc., Cary NC, USA) and R (Version 4.3.2; R Core Team, Vienna, Austria, 2023).

## RESULTS

Between July 2013 and June 2023, 5,339,989 in-hospital stays for LRTI were identified **(Table 1)**. Their median [Q1-Q3] age was 78 [65-87] years, and women accounted for 44.9% of inpatients. Overall, one third of patients (34.5%) had at least one CRD, mainly COPD (19.4%). Almost 25% of patients were admitted to the ICU during their hospital stay, 60.5% of whom directly. The median hospital length of stay was 9 [5-15] days, and 729,568 (13.7%) patients died.

**Table 1:** Characteristics of the population hospitalised for lower respiratory tract infection (excluding SARS-CoV-2 infection) in France from July 2013 to June 2023, overall and by study period.

|  | All study period,<br>N=5,339,989* | Pre-pandemic period,<br>July 2013 - March 2020<br>N=3,914,800* | Period 1,<br>April 2020 - May 2023<br>N=454,298* | Period 2,<br>June 2021 - June 2022<br>N=442,162* | Period 3,<br>July 2022 - June 2023<br>N=528,729* |
| --- | --- | --- | --- | --- | --- |
| **Age, years** | 78 [65-87] | 79 [65-87] | 76 [64-86] | 76 [64-86] | 76 [65-86] |
| **Female gender** | 2,397,253 (44.9%) | 1,778,552 (45.4%) | 186,733 (41.1%) | 192,220 (43.5%) | 239,748 (45.3%) |
| **CRD** | 1,842,925 (34.5%) | 1,351,657 (34.5%) | 153,779 (33.8%) | 153,690 (34.8%) | 183,799 (34.7%) |
| COPD | 1,033,930 (19.4%) | 771,619 (19.7%) | 80,799 (17.8%) | 82,931 (18.8%) | 98,581 (18.6%) |
| Asthma | 173,729 (3.3%) | 129,567 (3.3%) | 11,673 (2.6%) | 14,155 (3.2%) | 18,334 (3.5%) |
| CF-Bronchiectasis | 128,960 (2.4%) | 94,155 (2.4%) | 11,722 (2.6%) | 10,746 (2.4%) | 12,337 (2.3%) |
| ILD | 111,601 (2.1%) | 80,490 (2.1%) | 9,762 (2.1%) | 9,793 (2.2%) | 11,556 (2.2%) |
| Thoracic oncology | 235,558 (4.4%) | 163,889 (4.2%) | 24,963 (5.5%) | 21,955 (5.0%) | 24,751 (4.7%) |
| PVD | 129,683 (2.4%) | 91,194 (2.3%) | 12,491 (2.7%) | 12,098 (2.7%) | 13,900 (2.6%) |
| CRD associated with NMD | 8,410 (0.2%) | 6,265 (0.2%) | 719 (0.2%) | 654 (0.1%) | 772 (0.1%) |
| Lung transplantation | 12,473 (0.2%) | 8,677 (0.2%) | 1,310 (0.3%) | 1,181 (0.3%) | 1,305 (0.2%) |
| **Type of LRTI** |  |  |  |  |  |
| Pneumonia | 4,050,133 (75.8%) | 2,890,459 (73.8%) | 376,531 (82.9%) | 357,402 (80.8%) | 425,741 (80.5%) |
| AECOPD | 619,367 (11.6%) | 481,563 (12.3%) | 43,835 (9.6%) | 44,293 (10.0%) | 49,676 (9.4%) |
| AEBX | 22,644 (0.4%) | 16,729 (0.4%) | 2,035 (0.4%) | 1,890 (0.4%) | 1,990 (0.4%) |

|  | **All study period,** | **Pre-pandemic period,** | **Period 1,** | **Period 2,** | **Period 3,** |
| --- | --- | --- | --- | --- | --- |
|  |  | July 2013 - March 2020 | April 2020 - May 2023 | June 2021 - June 2022 | July 2022 - June 2023 |
|  | N=5,339,989* | N=3,914,800* | N=454,298* | N=442,162* | N=528,729* |
| Acute bronchitis | 647,845 (12.1%) | 526,049 (13.4%) | 31,897 (7.0%) | 38,577 (8.7%) | 51,322 (9.7%) |
| **Admission to the ICU** | 1,268,087 (23.7%) | 922,894 (23.6%) | 116,035 (25.5%) | 104,909 (23.7%) | 124,249 (23.5%) |
| Direct admission | 766,869 (60.5%) | 555,100 (60.1%) | 70,063 (60.4%) | 64,493 (61.5%) | 77,213 (62.1%) |
| Time to ICU admission, days | 0 [0-1] | 0 [0-1] | 0 [0-1] | 0 [0-1] | 0 [0-1] |
| Organ support | 795,779 (62.8%) | 581,481 (63.0%) | 72,290 (62.3%) | 64,737 (61.7%) | 77,271 (62.2%) |
| SAPSII | 33 [20-47] | 33 [21-47] | 32 [17-47] | 32 [18-47] | 33 [18-47] |
| **Hospital length of stay, days** | 9 [5-15] | 9 [5-15] | 9 [5-16] | 9 [4-15] | 8 [4-15] |
| **Hospital Death** | 729,568 (13.7%) | 511,725 (13.1%) | 74,118 (16.3%) | 67,935 (15.4%) | 75,790 (14.3%) |

\*Median [Q1-Q3]; n (%)
Abbreviations: CRD: chronic respiratory disease; COPD: chronic obstructive pulmonary disease; CF: cystic fibrosis; ILD: interstitial lung disease; PVD: pulmonary vascular disease; NDM: neuromuscular disease; LRTI: lower respiratory tract infection; AECOPD: acute exacerbation of COPD; AEBX: acute exacerbation of bronchiectasis; ICU: intensive care unit; SAPSII: simplified acute physiology score

Overall, the age of patients hospitalised decreased after the onset of the pandemic and remained stable thereafter: 79 [65-87] years in pre-COVID period and around 76 [64-86] years for the 3 periods. The proportion of women decreased after the start of the pandemic, followed by an increase in periods 2 and 3. Similarly, in period 1, there was a drop in the proportion of patients with CRD (33.8% versus 34.5% in pre-pandemic period), mainly due to a reduction in COPD and asthma. In addition, the number of deaths increased during the early stages of the pandemic from 13.1% to 16.3% at period 1, as did the number of ICU admissions (23.6% to 25.5%). Noteworthy, mortality among patients admitted to the ICU increased from 20.8% to 22% during period 1. Overall, 20.9% of patients with an ICU stay died over the entire study period.

A segmented regression model with autocorrelated error fulfilling goodness-of-fit criteria was obtained for each series (**Figure 1 and Figures S1-2, 4-6, 8-9, Supplementary data**). The estimated differences in monthly rates of hospitalisations per 100,000 inhabitants during the periods are reported for all the considered series in **Table S5 (Supplementary data)** (in incidence rates) and **Figures 2 and Figure S3 in Supplementary data** (in percent change relative to the pre-pandemic period).

**Figure 1:**
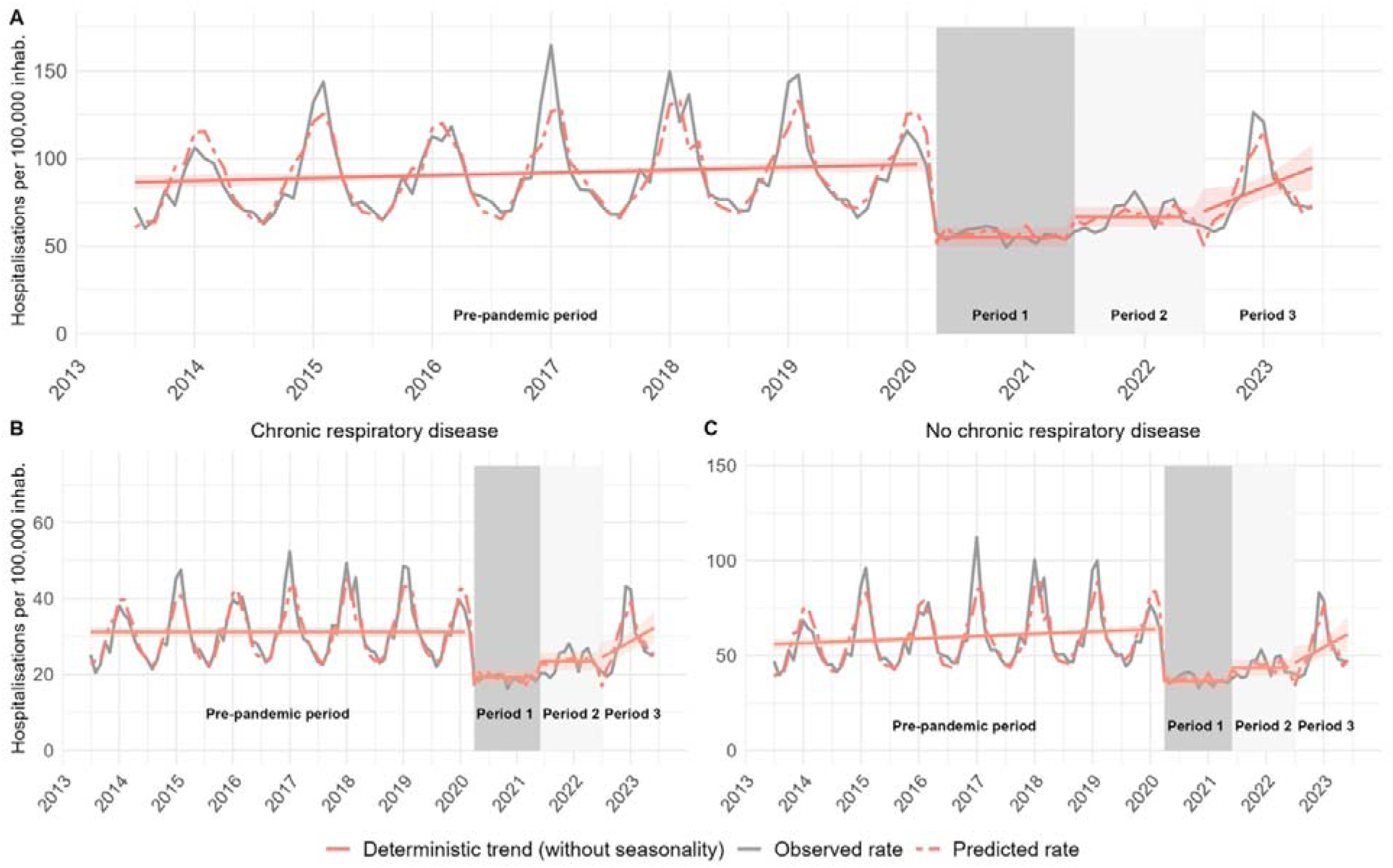
Monthly hospitalisation rates of lower respiratory tract infection (excluding COVID-19 infection). Presentation of all hospital admissions for LRTI (A), and according to the presence (B) or absence (C) of a CRD. The observed incidence is depicted by the grey line. The model prediction is presented by the dotted red line, the predicted deterministic trend (regression line) and its 95% confidence intervals by the solid red line.

**Figure 2:**
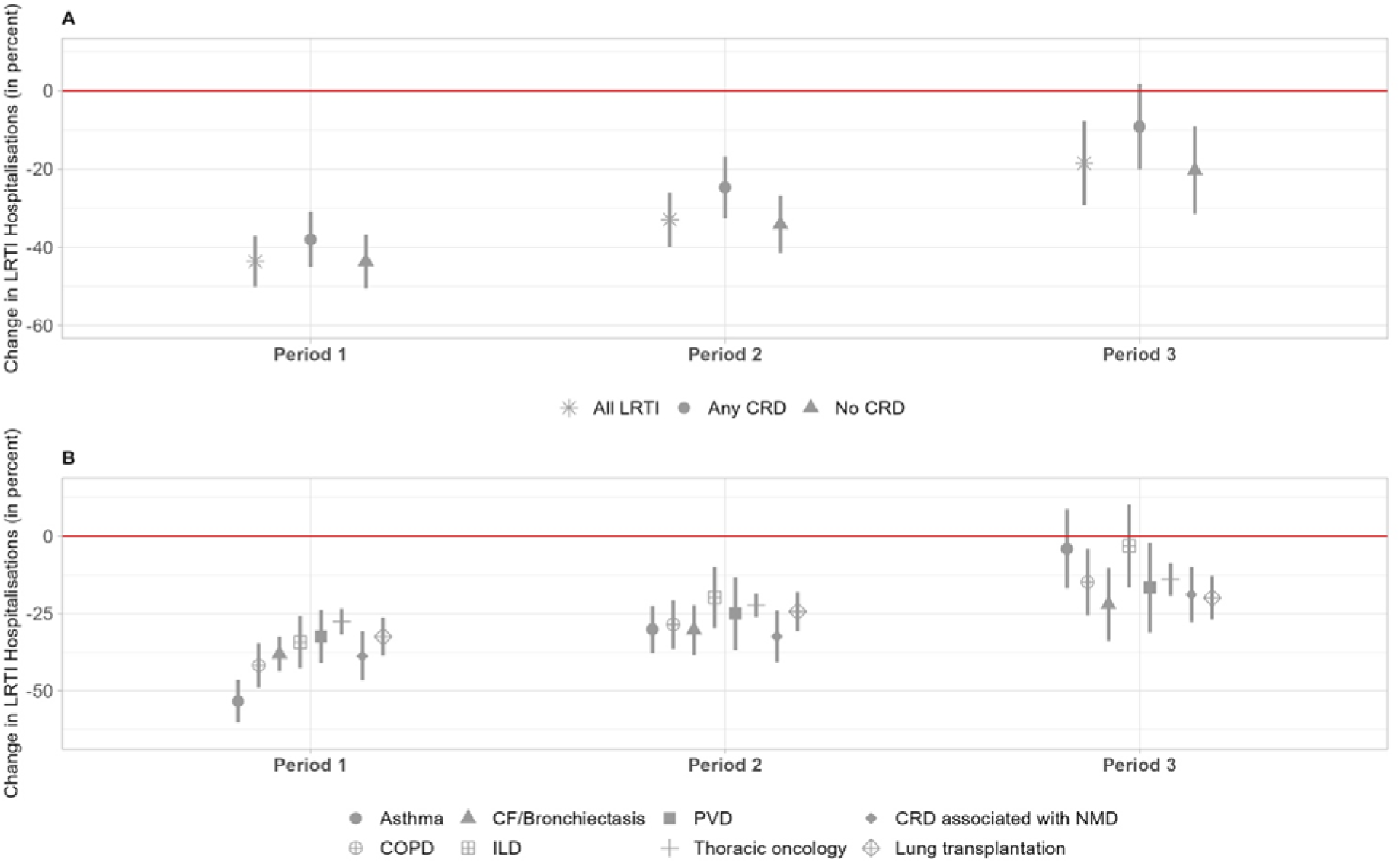
Change in LRTI hospitalisation rates during study periods according to the absence or presence of a CRD (A) and according to each category of CRD (B). Period 1 extends from April 2020 to May 2021, period 2 from June 2021 to June 2022 and period 3 from July 2022 to June 2023. Abbreviations: LRTI: Lower respiratory tract infection; CRD: Chronic respiratory disease; CF: cystic fibrosis; ILD: interstitial lung disease; PVD: pulmonary vascular disease; NMD: neuromuscular disease.

Monthly rate of hospitalisations for LRTI showed a winter seasonal pattern in the adult population during the pre-pandemic period, with a rising trend from 2013 to 2020 **(Figure 1A)**. Compared with the pre-pandemic period, a significant decrease in LRTI hospitalisations was observed in the early period 1 (−43.6%; 95% CI: −50.1% to −37.2%. **Figure 2A)**. This decline was associated with a loss of the seasonality pattern and slope. Compared with the pre-pandemic period, the decrease was maintained but less marked during period 2 (−33.0%; 95% CI: −39.9% to −26.1%), and even less pronounced during period 3 (−18.5%; 95% CI: - 29.2% to −7.9%). This was also associated with a significantly higher slope than in the pre-pandemic period (+2.3; 95% CI: 0.2 to 4.4; p=0.04) and a return to the seasonal pattern seen previously **(Figure 1A)**. The same overall tendency occurred in the presence of at least one CRD, with a significant decrease in LRTI hospitalisations when successive lockdown and strict NPI measures were implemented, followed by a gradual increase at periods 2 and 3 **(Figures 1B and 2A)**. Notably, during period 3, there was a resurgence of seasonality and an upward slope (+0.7; 95% CI: +0.1 to +1.4; p=0.04). Furthermore, this trend had almost returned to the pre-pandemic level during period 3 and was not statistically different (−9.2%; 95% CI: −20.9% to 1.7%). It should be noted that this reduction of hospitalisations in CRD inpatients was less pronounced than in those without CRD **(Figures 1C and 2A)**.

Focusing on the different categories of CRD, the drop of LRTI hospitalisations involved all respiratory diseases during period 1 **(Figure 2B and Figure S1, Supplementary data)**, and particularly asthma (−53.4%; 95% CI: −60.3% to −46.5%) and COPD (−41.8%; 95% CI: - 49.0% to −34.6%). At periods 2 and 3, there was a gradual reduction of these differences in all categories of CRD. Altogether, at period 3, the average level of LRTI hospitalisations did not return to the pre-pandemic level for COPD (−14.8%; 95% CI: −25.5% to −4.1%), CF/bronchiectasis (−22.0%; 95% CI: −33.8% to −10.2%), thoracic oncology (−13.9%; 95% CI: −19.3% to −8.6%), lung transplantation (−19.9%; 95% CI: −26.9% to −12.8%) and CRD associated with NMD (−18.8%; 95% CI: −27.8% to −9.8%) **(Figure S1, Supplementary data)**. Conversely, this difference was no longer significant for asthma (−4.1%; 95% CI: - 16.8% to +8.7%) and ILD (−3.1%; 95% CI: −16.5% to +10.4%), as compared with the pre-pandemic period.

The decline in LRTI hospitalisations was more pronounced for moderately severe LRTI, as compared with severe LRTI and those associated with death **(Figures S2 and S3A, Supplementary data)**. In addition, moderate forms showed a significant upward trend during study period 3 (+1.8; 95% CI: +0.2 to +3.3; p=0.03), a trend not seen in the two other forms **(Table S6 and Figure S3A, Supplementary data)**. Notably, at period 3, the decrease in LRTI hospitalisations did not differ from that of the pre-pandemic period for the episodes associated with death (−2.2%; 95% CI: −10.9% to +6.4%).

Focusing on pneumonia, a sharper decline occurred during period 1 (−36.3%; 95% CI: −42.9% to −29.7%) than period 2 (−27.6%; 95% CI: −34.3% to −21.0%) **(Figures S3B and S4A, Supplementary data)**. In period 3, there was a lower but still significant decrease (−13.3%; 95% CI: −22.9% to −3.8%). For other types of LRTI, more pronounced trends were observed with a respective decrease of 56.6% (95% CI: −68.4% to −44.7%) at period 1, 45.7% (95% CI: −58.1% to −33.4%) at period 2 and 32.7% (95% CI: −45.6% to −19.9%) at period 3 **(Figure S3B and S4B, Supplementary data)**. Moreover, unlike pneumonia, there was no increasing slope during period 3, although there was a seasonality pattern.

Although the decline in hospitalisations was greater among women, there was a marked decrease in period 1 in both genders. This change was gradually less important during periods 2 and 3 compared to the pre-pandemic period **(Figure S3C and Figure S5, Supplementary data)**. This feature was similar for all age categories studied, although the decline was greatest among oldest patients (i.e., over 75 years) **(Figure S3D and S6, Supplementary data)**.

Concurrent with the increase in LRTI hospitalisations, the proportion of COVID-19 hospitalisations among all LRTI decreased from period 1 to period 3 (**Figure S7, Supplementary data**). During period 1, they accounted for nearly half of all respiratory infections hospitalised, dropping to one-third during period 2, and less than 20% during the last epidemic season despite the reduction of NPI **(Table S7, Supplementary data)**. Interestingly, COVID-19 hospitalisations exhibited a winter seasonality comparable to other hospitalised LRTI at period 3. In this context, when comparing period 3 with the pre-pandemic period, the overall burden of hospitalisations for LRTI, (including COVID-19), is comparable to that of the pre-pandemic period (+0.3; 95% CI: −9.0% to +9.7%) (**Figure S8 and Table S6, Supplementary data**). Notably, when a CRD is present, the increase in hospitalisations is more important at period 3 compared with the predicted level **(Figure S9, Supplementary data)**, but not statistically significant: + 6.1% (95% CI: −3.4% to +15.7%) versus +1.0% (95% CI-8.9% to +11.0%) if no CRD is present.

## DISCUSSION

In this nationwide study using The French National Hospital Discharge Database, we showed a marked decrease of hospital admissions for LRTI unrelated to COVID-19 at the onset of the pandemic, followed by a subsequent rise coinciding with the lifting of COVID-19 control measures and a return to a seasonal pattern, particularly notable in patients with CRD. This increase in LRTI hospitalisations was associated with a return to pre-pandemic levels within this group, with some discrepancies depending on the type of CRD.

Our findings document the significant decline in hospital admissions for LRTI unrelated to COVID-19 when NPI measures were implemented to control the pandemic. This trend has been well documented in the literature, as these measures reduced the circulation of respiratory viruses (22–24), as well as some bacteria typically associated with LRTI (10,25). Following the partial relaxation and eventual complete lifting of COVID-19 control measures, a notable resurgence in hospitalisations for other LRTI was observed. This trend has been previously demonstrated, specifically in the paediatric population, and coincided with the sustained resurgence of the RSV epidemic (26). While post-pandemic COVID-19 data in the adult population are lacking, several studies have indicated an increase in the incidence of other LRTI following the easing of restrictions (23,27,28). Our study further contributes to this understanding, highlighting that this increase is not only important when NPI are relaxed, but also follows a typical winter seasonal pattern consistent with pre-pandemic trends, underlining the role of seasonal viruses. Community-acquired pneumonia exhibited a lesser reduction in hospitalisations compared to non-pneumonic LRTI, such as exacerbations of COPD, bronchiolitis, and acute bronchitis. Pneumonia is indeed recognized as the primary cause of hospitalisations among all LRTI (29). The frequent involvement of respiratory viruses during acute bronchitis (30) and acute exacerbations of COPD (31) may have been particularly affected by NPI. However, during the period when NPI were completely lifted, the resurgence of non-pneumonic LRTI was less significant than that of pneumonia. This could be attributed to an increased focus on outpatient management of non-pneumonic LRTI during and extending after the pandemic. Furthermore, the continued adherence to some NPI by vulnerable populations (32) and health professionals (33) may have contributed to mitigating the increase in LRTI incidence beyond the pandemic.

Changes in hospital admissions for LRTI were primarily driven by mild-to-moderately severe forms of LRTI. Indeed, our findings indicate that the decrease in episodes of LRTI leading to ICU admission or death was less pronounced than that observed for mild-to-moderately severe forms of LRTI. It can be assumed that some of the latter cases were managed through outpatient care or required fewer medical consultations (34,35). Additionally, while the overall level of LRTI hospitalisations may not be entirely comparable to that of the pre-pandemic period, the rate of fatal LRTI episodes remained similar. This suggests that the criteria for hospitalisation for LRTI may have been more stringent and that hospitalised patients may have been more severe.

We found that patients with CRD accounted for 30% of all LRTI inpatients, with half of them diagnosed with COPD. Our findings underscore the significant impact of NPI on patients with CRD, demonstrating a substantial decrease in hospitalisations for LRTI during the most restrictive NPI period, followed by a subsequent rise during the relaxation phases. Interestingly, a return to pre-pandemic levels was observed in CRD patients upon complete lifting of restrictions. This observation is particularly interesting as it highlights the vulnerability of this population to LRTI, while also emphasizing the critical role of NPI in mitigating the burden of LRTI among CRD individuals (23,36). This observation may be partly supported by a reduction in the circulation of respiratory viruses during the pandemic, a factor well-documented to trigger CRD exacerbations, notably asthma and COPD (23). Indeed, we observed the most significant initial decline in hospitalisations for these two populations. Moreover, we noted a return to pre-pandemic levels among hospitalised asthma patients. In contrast, for COPD patients, who accounted for more than half of CRD hospitalisations for LRTI, a notable increase was observed after removal of NPI, although it did not reach pre-pandemic levels. One possible explanation for this partial return to pre-pandemic levels might be an ongoing adherence to protective measures, such as wearing masks in the community and systematic hand washing (37). Interestingly, patients with ILD experienced a significant increase in hospitalisations following the lifting of NPIs, returning to pre-pandemic levels. LRTI account for approximately 20% of admissions in these patients (38) and may worsen their condition (39), emphasizing the importance of maintaining NPI measures in this population.

Following the complete release of NPI, COVID-19 still accounted for 20% of hospitalised LRTI, and exhibited an epidemic pattern overlapping with other LRTI, highlighting the propensity for SARS-CoV-2 to become seasonal (18). Despite variations in the pathogens involved, the overall level of LRTIs in period 3 did not significantly surpass pre-pandemic levels, indicating that the simultaneous occurrence of multiple respiratory pathogen epidemics did not lead to an excessive increase in hospitalisations for LRTIs in France. Finally, it is noteworthy that CRD patients experienced a more significant increase, highlighting the particularly vulnerability of this population to respiratory pathogens, including COVID-19 (40).

Our study has several strengths. Firstly, it utilizes national data encompassing all hospitalisations in France over a 10-year period. Secondly, the study period covers a complete epidemic season after the end of all NPI recommendations, allowing an examination of overall trends in hospitalisations for LRTI. Moreover, our approach provides an estimate of the burden of LRTI in patients with CRD at the national level post-pandemic. Identifying key interventions for LRTI control and prospectively demonstrating which of these are most acceptable and easily followed by vulnerable populations, such as CRD patients, could be effective in reducing the burden of these conditions on hospitals. While implementing all NPI measures like lockdowns may not be feasible or acceptable, some could be beneficial, particularly among patients with CRD. For example, measures such as mask-wearing have been shown to have a real impact on reducing the circulation of certain viruses such as RSV (22). Moreover, some of these less restrictive measures appear to be adhered to among patients with CRD (32,37). Therefore, targeted implementation of specific NPI on vulnerable populations (i.e., CRD, as well as older individuals) could be considered as a viable strategy for mitigating the burden of respiratory infections.

Our study presents limitations. Although our study suggests that the lifting of NPI impacts hospital admissions for LRTI, we were unable to determine which measures have the most significant impact. Furthermore, this is an observational study investigating the temporal relationship between hospitalisations for LRTI and NPI, but we cannot establish a strict correlation and cannot exclude the possibility that other factors may have influenced these variations. Moreover, medico-administrative databases have restricted access to clinical data, which limits the accuracy of our diagnoses. This limitation is inherent to such databases, and we utilized ICD-10 codes commonly used for research in these disease categories.

Nevertheless, the French public health insurance system regularly audits and randomly checks that coding records correspond to actual patient records, thereby reducing coding errors (41). Finally, due to imperfect pathogen coding in these databases, with many LRTI remaining unidentified, we could only study LRTI collectively.

Our study emphasizes that the lifting of restrictions is associated with a rapid return to the typical seasonality of LRTI. Hospitalisations for LRTI among patients with CRD returned to pre-pandemic levels. Finally, given the persistent circulation of COVID-19, its influence on hospitalisations remains significant.

## Data Availability

Aggregated monthly data were used for the time series analysis and are available from the corresponding author on reasonable request. The pseudonymized data extracted from the PMSI are only accessible to accredited structures and can be accessed with the permission from the Agence Technique de l Information sur l Hospitalisation

## Aknowledgment

The authors thank Bérénice Varga for her careful revision of the manuscript.

## Funding information

Doctoral fellowship was received by ASE from Université Paris-Saclay. The funder has no role in the conceptualization, design, data collection, analysis, decision to publish, or preparation of the manuscript.

## Competing Interests

ASE reports support for attending meeting from SOS Oxygène and LVL Medical outside the submitted work. MF reports grants from BioMérieux and personal fees from BioMérieux, Pfizer, SOS Oxygène and Fisher and Paykel outside the submitted work. LW reports personal fees from Pfizer, Sanofi and Heva outside the submitted work. DG, CBB and LB have no competing interests to declare.

## Notes

### Competing Interest Statement

ASE reports support for attending meeting from SOS Oxygene and LVL Medical outside the submitted work. MF reports grants from BioMerieux and personal fees from BioMerieux, Pfizer, SOS Oxygene and Fisher and Paykel outside the submitted work. LW reports personal fees from Pfizer, Sanofi and Heva outside the submitted work. DG, CBB and LB have no competing interests to declare.

### Funding Statement

Doctoral fellowship was received by AES from Paris-Saclay University. The funder has no role in the conceptualization, design, data collection, analysis, decision to publish, or preparation of the manuscript.

### Author Declarations

The study was conducted using the Institut National de la Sante et de la Recherche Medicale permanent access to the the French National Hospital Discharge Database.

