## Supplementary data for "Trends in hospitalisations for lower respiratory tract infection after the COVID-19 pandemic in adults with chronic respiratory disease"

### SUPPLEMENTARY METHODS

### Data source:

The French National Hospital Discharge Database (PMSI: Programme de Médicalisation des Sytèmes d’Informations) covers the activity of all public and private health care facilities in France with approximately 12.6 million hospitalisations over the 2017-2021 period (1). For each hospitalisation, the cause for hospital admission is specified by International Classification of Disease (10^th^ revision) codes detailed by a principal diagnosis (PD: condition requiring hospitalisation), related diagnosis (RD: adds information to PD) and one or more secondary associated diagnoses (SAD: complications and co-morbidities potentially affecting the course or cost of hospitalisation). Demographic data were obtained from the French National Institute of Statistics and Economic Studies (<https://www.insee.fr/fr/accueil>).

LRTI was defined by the presence of an ICD-10 code as PD or SAD during hospital stay (**Table S1**).

**Table S1 : ICD-10 codes for the selection of patient with LRTI and COVID-19**

| **LRTI classification** | **ICD-10 code^*^** |
| --- | --- |
| Pneumonia | A48.1, B20.6, B48.5^†^, B59^⁋^, J10.0, J11.0, J12.0, J12.1, J12.2, J12.3, J12.8, J12.9, J13, J14, J15.0, J15.1, J15.2, J15.3, J15.4, J15.5, J15.6, J15.7, J15.8, J15.9, J16.0, J17.0, J17.1, J17.2, J17.3, J17.8, J18.0, J18.1, J18.2, J18.9, J69.0, J85.0, J85.1, J85.2, J86.0, J86.9, J88.8 |
| COVID-19^§^ | U07.10, U07.11, U10.9 |
| Other LRTI | J09, J10.1, J11.1, J20.1, J20.2, J20.3, J20.4, J20.5, J20.6, J20.7, J21.0, J22, J20.8, J20.9, J44.0 |

**^*^**ICD-10 codes were used to define LRTI diagnoses in Principal Diagnosis or in Secondary associated diagnoses. ^†^Used in the ICD-10 classification after 2019; ^⁋^Used in the ICD-10 classification before 2019. ^§^If these codes are missing during hospitalisation, the patient is classified as LRTI not associated with COVID-19.

Abbreviation: LRTI: Lower respiratory tract infection; ICD-10: International Classification of Disease – 10^th^ revision.

To evaluate the level of disease severity, we defined 3 groups according to the evolution during hospitalisation: (i) death, (ii) severe: intensive care unit admission and/or organ support, (iii) moderate. Severe evolution was determined by the presence of a Classification Commune des Actes Médicaux (CCAM) code associated with organ failure or intensive care hospitalisation (Medical Unit code). This classification was based on recent recommendations in management of severe community-acquired pneumonia (2). Otherwise, the disease was defined as moderate.

**Table S2 : CCAM codes and medical unit codes (UM) used to define severe conditions of LRTI**

| UM codes from PMSI^*^ | 01A, 02A, 02B, 03A, 18 |
| --- | --- |
| CCAM codes^†^ | GLLD003, GLLD004, GLLD008, GLLD012, GLLD015, GLLD019, EQLF002, EQLF003, DKMD001, DKMD002, JVJB002, JVJF002, JVJF003, JVJF005 |

^*^ UM codes are used to define the hospital care department during the stay; ^†^CCAM codes are used to determine the presence of organ support during the stay.

Abbreviations: UM : Unité médicale (Medical unit); PMSI : Programme de Médicalisation des Sytèmes d’Informations (French National Hospital Discharge Database) ; CCAM : Classification Commune des Actes Médicaux (the French classification of Medical Procedures).

**Table S3: ICD-10 codes used to define chronic respiratory disease**

| **Chronic respiratory disease classification** | **ICD-10 code^*^** |
| --- | --- |
| Chronic non-interstitial lung disease | J41, J42, J43, J44 |
| Asthma | J45, J46 |
| Cystic fibrosis/Other bronchiectasis | E840, J47 |
| Chronic respiratory failure associated with neuromuscular disease | G12 + J99 or J96.1 or J96.9  G35 + J99 or J96.1 or J96.9  G37 + J99 or J96.1 or J96.9  G70 + J99 or J96.1 or J96.9  G71 + J99 or J96.1 or J96.9  G73 + J99 or J96.1 or J96.9 |
| Interstitial lung disease | J60, J61, J62, J63, J64, J65, J66, J67, J68.4, J70.1, J70.3, J82, J84, J99 |
| Thoracic oncology | C33, C34, C38, C39, C45.0, Z85.1 |
| Pulmonary vascular disease | I27.0, I27.2, I27.9, I28.9  I78.0 + J99 or J96.1 or J96.9 |
| Lung transplantation | Z94.2, Z94.3 |
| Others CRD | B90.9, G47.3, E66.2, J92, J95.3, J96.1, J96.9, J98.0, J98.2, J98.3, J98.4, J98.5, J98.6, J98.9 |

^*^ICD-10 codes were used to define CRD diagnoses in Principal Diagnosis or in Secondary associated diagnoses.

Abbreviation: ICD-10: International Classification of Disease – 10^th^ revision.

**Table S4: Main non-pharmaceutical interventions implemented in France and key vaccination dates during the COVID-19 pandemic by study period**

| **Period 1: April 2020 to May 2021** | **17 March 2020 to 11 May 2020** | **Stay at home order.**  Total closure of primary, secondary schools and Universities. |
| --- | --- | --- |
|  | 6 June 2020 | Restaurant opening only outside. |
|  | 16 June 2020 | Partial reopening of primary and secondary schools. |
|  | 22 June 2020 | Opening of cinema, theatre. |
|  | 11 July 2020 | Opening of stadiums and major events with a maximum capacity of 5,000 people. |
|  | 18 August 2020 | Masks indoor mandatory. |
|  | 1^st^ September 2020 | Back-to-school in all establishments possible   - Mask mandatory. - Class closure after 3 COVID-19 positive cases. |
|  | From 23 September to 13 October 2020 | Gradual closure of bars across the country. |
|  | From 17 October to 22 October 2020 | Gradual nationwide extension of curfews (9 p.m. to 6 a.m.). |
|  | **30 October 2020 to 15 December 2020** | **Stay at home order.**  Schools remain open during lockdown. |
|  | 15 December 2020 | Curfew (8 p.m. to 6 a.m.). |
|  | 27 December 2020 | Start of the COVID-19 vaccination campaign |
|  | 16 January 2021 | Curfew extended from 6 p.m. to 6 a.m. |
|  | **3 April 2021 to 3 May 2021** | **Stay at home order.**  Total closure of primary, secondary schools and Universities until April 26.  Mask mandatory indoor and outdoor at the end of the lockdown. |
|  | 19 May 2021 | Curfew eased (starting at 9 p.m.).  Restaurant opening only outside at 50% of capacity.  Open cinema, theater, and indoor sports facilities at 35% of capacity. |
| **Period 2: June 2021 to June 2022** | 9 June 2021 | **Sanitary pass effective date,** valid if   - Complete vaccination schedule. - COVID-19 test negative for less than 24h. - Previously proven COVID-19 recovery.   Inside restaurant/bar and indoor sport facilities capacity at 50% and cinema/theater at 65%.  Curfew eased (starting at 11 p.m.).  End of the full-time teleworking |
|  | 17 June 2021 | Mask not mandatory outdoors. |
|  | 20 June 2021 | End of curfew. |
|  | 27 June 2021 | One third of the population with full vaccination schedule. |
|  | 30 June 2021 | End of outdoor limit gauges.  Complete restaurant reopening.  Complete reopening of cinema, theater, and cultural events (with possible local restrictions). |
|  | 9 July 2021 | Nightclub reopening. |
|  | 1^st^ August 2021 | 50 percent of the population with full vaccination schedule. |
|  | 5 September 2021 | Two thirds of the population with full vaccination schedule. |
|  | 10 December 2022 | Nightclub closures. |
|  | 3 January 2022 | Mandatory telecommuting 3 days a week.  Maximum capacity for major events: 2,000 people indoors, 5,000 outdoors. |
|  | 2 February 2022 | Telecommuting not mandatory.  Masks in outdoor public spaces no longer mandatory.  Reception capacity gauges are removed in establishments open to the public (only seated). |
|  | 16 February 2022 | Nightclub reopening.  End of limitation on non-sitting concerts. |
|  | 18 February 2022 | Mask no longer mandatory in enclosed public areas. |
|  | 14 March 2022 | End of sanitary/vaccination pass (except in healthcare establishments). |
| **Period 3: to August 2022 to June 2023** | 1^st^ August 2022 | End of all exceptional measures concerning the COVID-19 pandemic. |

### Mathematical formulation of the model and estimated changes:

Each time series was studied using a regression model with autocorrelated errors. The segmented regression model can be written as follow:

$$Y_{t}=\sum_{i=0}^{3} \left( \beta_{0i}+\beta_{1i}t \right)I_{i}+\sum_{i=0,3} \left( \gamma_{i}cos\frac{2\pi_{t}}{12}+\delta_{i}sin\frac{2\pi_{t}}{12} \right) I_{i}+\nu_{t}$$

Notation:

t = time index, from 1 to 120,

i = period index, from 0 to 3, i=0 before the pandemic period (July 2013 to March 2020),

$Y_{t}$ = number of hospitalisations of interest at time t (month) per 100,000 inhabitants,

$I_{i}$ = dummy variable for period i,

$\beta_{0i}$= intercept parameter for period i,

$\beta_{1i}=$slope parameter for period i,

$\gamma_{i}$ and $\delta_{i}$ = cosinus and sinus parameters of period i,

$\nu_{t}$ is modelled as an AR(p) process, with residual variance $\sigma^{2}.$

For each period, a regression line with a trigonometric function, if the series exhibited annual seasonality, was estimated. When non-significant, parameters were removed from the model.

To assess the validity of the model, Ljung-Box test was used to test if residuals were independently distributed and Shapiro-Wilk normality test for the Gaussian distribution of the residuals.

From the model, for each period i (i=1 to 3) and $t^{*}$ is the middle of the interval, the estimated level is defined by $\hat{Y}_{t^{*}i}=\beta_{0i}+\beta_{1i}t^{*}$ and the predicted one, under the assumption that no change in the evolution, is $\overline{Y}_{t^{*}}=\beta_{00}+\beta_{10}t^{*}$. Afterwards, the subscripts are suppressed in order not to overburden the notation.

For each period, absolute difference was defined by:

$$AD=\hat{Y}-\bar{Y}$$

and percentage change by:

$PC=\frac{\hat{Y}-\overline{Y}}{\hat{Y}}\times100$*.*

In cases where a difference in slope was observed before pandemic and subsequent periods, estimations were calculated at the mid-point of considered subsequent intervals.

If variance of AD can be obtained by using different scripts (*e.g.* by estimating an intercept over the entire studied period), a delta method approach, as described by Zhang *et al.* (3), was used to approach the variance of PC.

The variance of $100\times PC$ was defined as follows:

$$Var\left( \frac{\hat{Y}-\overline{Y}}{\overline{Y}} \right)=\left( \frac{\hat{Y}}{\overline{Y}} \right)^{2}\left[ \frac{Var\left( \hat{Y} \right)}{\hat{Y}^{2}}+\frac{Var\left( \overline{Y} \right)}{\overline{Y}^{2}}-2\frac{Cov\left( \hat{Y},\overline{Y} \right)}{\hat{Y}\overline{Y}} \right]$$

Then 95% confidence interval can be calculated.

### SUPPLEMENTARY RESULTS

### Table S5: Estimated variation in hospitalisation rates of lower respiratory tract infections by epidemiological period

|  | **Period 1** | | **Period 2** | | **Period 3** | |
| --- | --- | --- | --- | --- | --- | --- |
|  | **Expected level,** | **Estimated difference,** | **Expected level,** | **Estimated difference,** | **Expected level,** | **Estimated difference,** |
|  | per 100,000 inh. (95%CI) | per 100,000 inh. (95%CI) | per 100,000 inh. (95%CI) | per 100,000 inh. (95%CI) | per 100,000 inh. (95%CI) | per 100,000 inh. (95%CI) |
| **All LRTI** | 97.67  (92.97 to 102.37) | -42.62  (-50.23 to -35.02) | 99.39  (93.57 to 105.21) | -32.77  (-40.86 to -24.68) | 100.92  (94.09 to 107.75) | -18.65  (-30.11 to -7.19) |
| **Severity** |  |  |  |  |  |  |
| Deaths | 11.83  (11.23 to 12.43) | -2.72  (-3.57 to -1.87) | 11.83  (11.23 to 12.43) | -1.53  (-2.47 to -0.59) | 11.83  (11.23 to 12.43) | -0.26  (-1.3 to 0.77) |
| Severe forms | 17.59  (16.85 to 18.34) | -5.66  (-6.91 to -4.41) | 17.59  (16.85 to 18.34) | -4.55  (-5.99 to -3.11) | 17.59  (16.85 to 18.34) | -1.90  (-3.42 to -0.39) |
| Moderate forms | 65.9  (62.39 to 69.41) | -31.53  (-37.21 to -25.86) | 67.17  (62.83 to 71.52) | -23.86  (-29.91 to -17.81) | 68.3  (63.2 to 73.4) | -13.85  (-22.43 to -5.26) |
| **Type of LRTI** |  |  |  |  |  |  |
| Pneumonia | 73.07  (69.6 to 76.55) | -26.54  (-32.2 to -20.89) | 74.66  (70.36 to 78.95) | -20.6  (-26.38 to -14.83) | 76.07  (71.03 to 81.1) | -10.11  (-17.79 to -2.42) |
| Other LRTI | 23.64  (22.31 to 24.96) | -13.36  (-16.37 to -10.35) | 23.64  (22.31 to 24.96) | -10.81  (-13.93 to -7.69) | 23.64  (22.31 to 24.96) | -7.74  (-10.93 to -4.54) |
| **Gender** |  |  |  |  |  |  |
| Female | 85.49  (80.7 to 90.28) | -42.28  (-50.03 to -34.53) | 87.12  (81.2 to 93.05) | -31.97  (-40.22 to -23.73) | 88.57  (81.61 to 95.54) | -16.78  (-28.5 to -5.06) |
| Male | 110.4  (105.39 to 115.42) | -42.21  (-49.57 to -34.85) | 112.13  (105.96 to 118.30) | -32.22  (-40.37 to -24.08) | 113.67  (106.44 to 120.89) | -20.06  (-31.67 to -8.45) |
| **Age** |  |  |  |  |  |  |
| 15-54 years | 15.74  (15.28 to 16.2) | -6.28  (-7.02 to -5.54) | 15.97  (15.4 to 16.54) | -4.34  (-5.14 to -3.53) | 16.17  (15.51 to 16.84) | -1.61  (-2.94 to -0.28) |
| 55-64 years | 62.39  (60.04 to 64.74) | -23.77  (-27.61 to -19.92) | 63.12  (60.21 to 66.02) | -17.35  (-21.44 to -13.26) | 63.76  (60.35 to 67.17) | -9.86  (-15.78 to -3.94) |
| 65-74 years | 138.75  (131.32 to 146.17) | -51.06  (-62.38 to -39.74) | 140.64  (131.49 to 149.79) | -38.55  (-50.64 to -26.46) | 142.31  (131.59 to 153.04) | -17.67  (-33.71 to -1.63) |
| >75 years | 462.38  (437.88 to 486.88) | -197.05  (-240.01 to -154.09) | 462.38  (437.88 to 486.88) | -149.27  (-194.94 to -103.60) | 462.38  (437.88 to 486.88) | -83.30  (-147 to -19.6) |
| **CRD** |  |  |  |  |  |  |
| Yes | 31.21  (29.95 to 32.47) | -11.86  (-14.19 to -9.54) | 31.21  (29.95 to 32.47) | -7.71  (-10.3 to -5.12) | 31.21  (29.95 to 32.47) | -2.87  (-6.31 to 0.56) |
| No | 64.70  (61.38 to 68.02) | -28.25  (-33.61 to -22.9) | 66.04  (61.93 to 70.15) | -22.56  (-28.26 to -16.86) | 67.23  (62.4 to 72.06) | -13.62  (-21.68 to -5.56) |
| **Type of CRD** |  |  |  |  |  |  |
| COPD | 17.82  (17.12 to 18.53) | -7.45  (-8.82 to -6.08) | 17.82  (17.12 to 18.53) | -5.08  (-6.57 to -3.6) | 17.82  (17.12 to 18.53) | -2.63  (-4.57 to -0.69) |
| Asthma | 3.02  (2.93 to 3.11) | -1.61  (-1.83 to -1.39) | 3.02  (2.93 to 3.11) | -0.91  (-1.15 to -0.67) | 3.02  (2.93 to 3.11) | -0.12  (-0.51 to 0.26) |
| CF/Bronchiectasis | 2.36  (2.12 to 2.59) | -0.9  (-1.07 to -0.72) | 2.41  (2.14 to 2.69) | -0.73  (-0.99 to -0.48) | 2.47  (2.15 to 2.79) | -0.54  (-0.88 to -0.2) |
| ILD | 1.85  (1.75 to 1.95) | -0.63  (-0.8 to -0.47) | 1.85  (1.75 to 1.95) | -0.36  (-0.56 to -0.17) | 1.85  (1.75 to 1.95) | -0.06  (-0.31 to 0.19) |
| PVD | 2.41  (2.14 to 2.68) | -0.78  (-1.04 to -0.52) | 2.5  (2.17 to 2.83) | -0.63  (-0.98 to -0.27) | 2.58  (2.2 to 2.96) | -0.43  (-0.86 to 0) |
| Thoracic oncology | 4.23  (4.11 to 4.34) | -1.17  (-1.36 to -0.97) | 4.34  (4.2 to 4.47) | -0.97  (-1.15 to -0.78) | 4.44  (4.27 to 4.60) | -0.62  (-0.87 to -0.37) |
| CRD associated with NMD | 0.15  (0.14 to 0.15) | -0.06  (-0.07 to -0.04) | 0.15  (0.14 to 0.15) | -0.05  (-0.06 to -0.03) | 0.15  (0.14 to 0.15) | -0.03  (-0.04 to -0.01) |
| Lung transplantation | 0.23  (0.22 to 0.24) | -0.08  (-0.09 to -0.06) | 0.24  (0.23 to 0.26) | -0.06  (-0.08 to -0.04) | 0.25  (0.24 to 0.27) | -0.05  (-0.07 to -0.03) |
| Abbreviations: LRTI: Lower respiratory tract infection; CRD: chronic respiratory disease; COPD: chronic obstructive pulmonary disease; CF: cystic fibrosis; ILD: interstitial lung disease; PVD: pulmonary vascular disease; NMD: neuromuscular disease; inh.: inhabitants | | | | | | |
| Period 1 extends from April 2020 to May 2021, period 2 from June 2021 to June 2022 and period 3 from July 2022 to June 2023. | | | | | | |

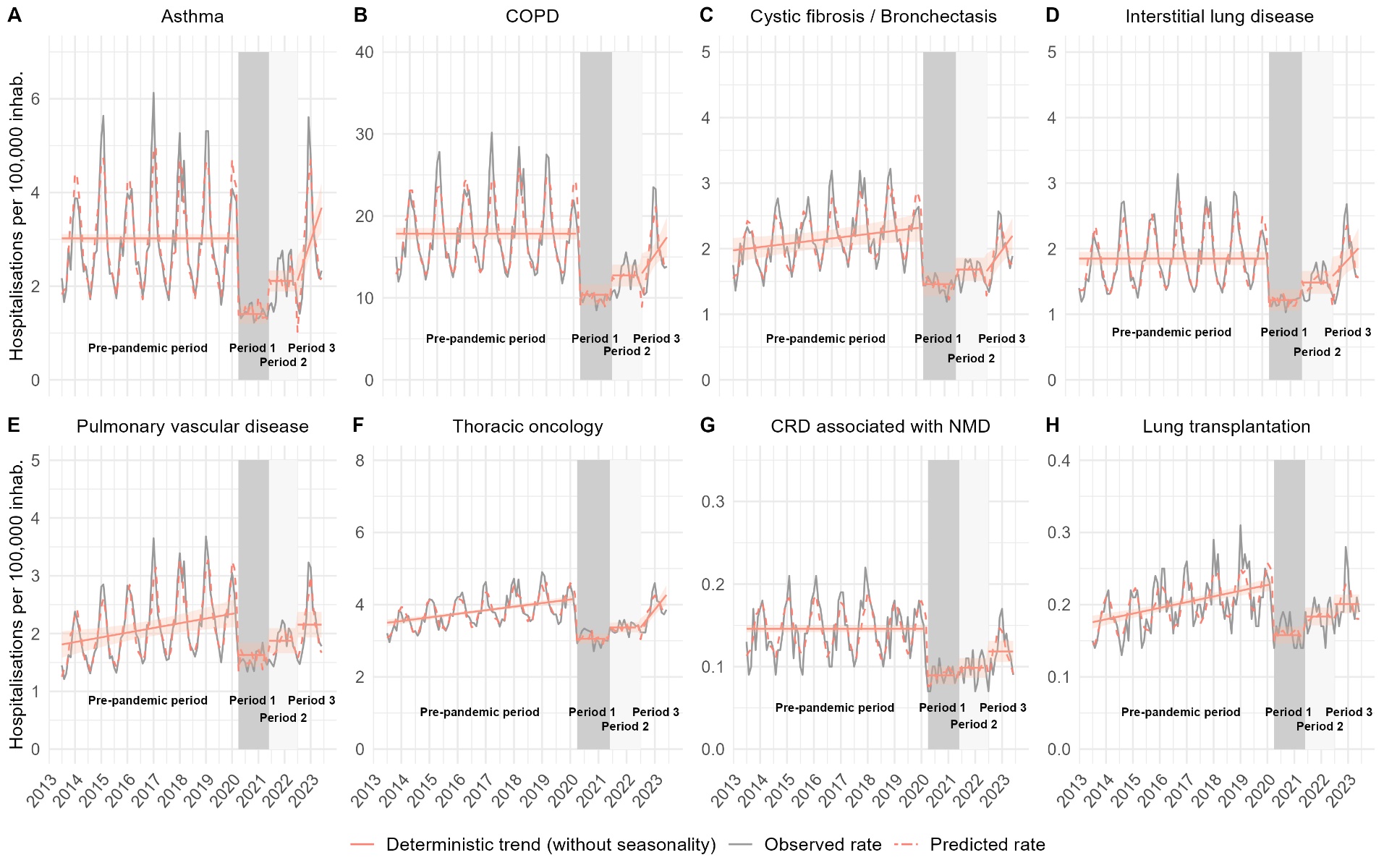

**Figure S1: Monthly hospitalisation rates of lower respiratory tract infection (excluding COVID-19 infection)** **by type of chronic respiratory disease.** The observed incidence is depicted by the grey line. The model prediction is presented by the dotted red line, the predicted deterministic trend (regression line) and its 95% confidence intervals by the solid red line.

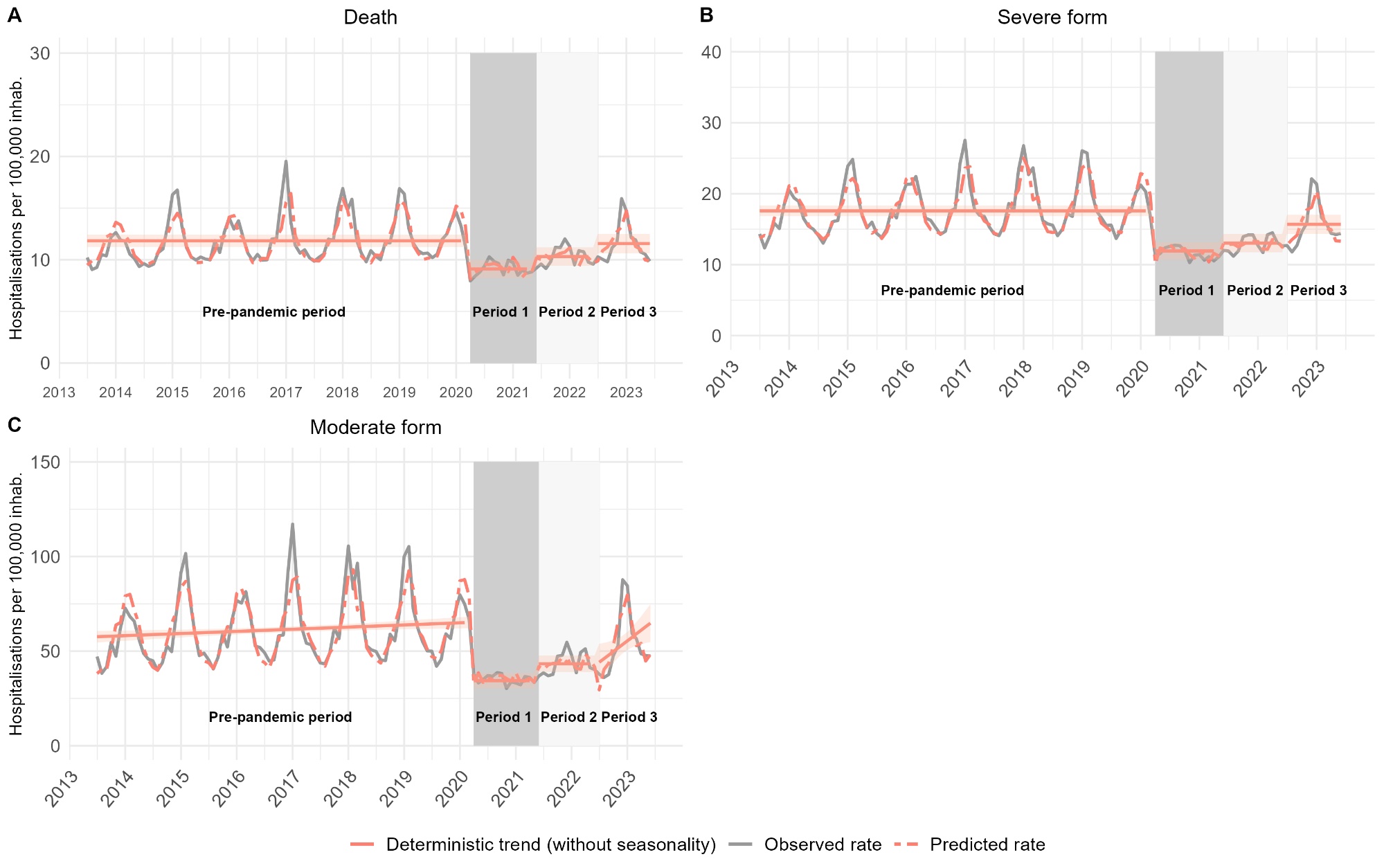

**Figure S2 Monthly hospitalisation rates of lower respiratory tract infection according to clinical severity**

The observed incidence is depicted by the grey line. The model prediction is presented by the dotted red line, the predicted deterministic trend (regression line) and its 95% confidence intervals by the solid red line.

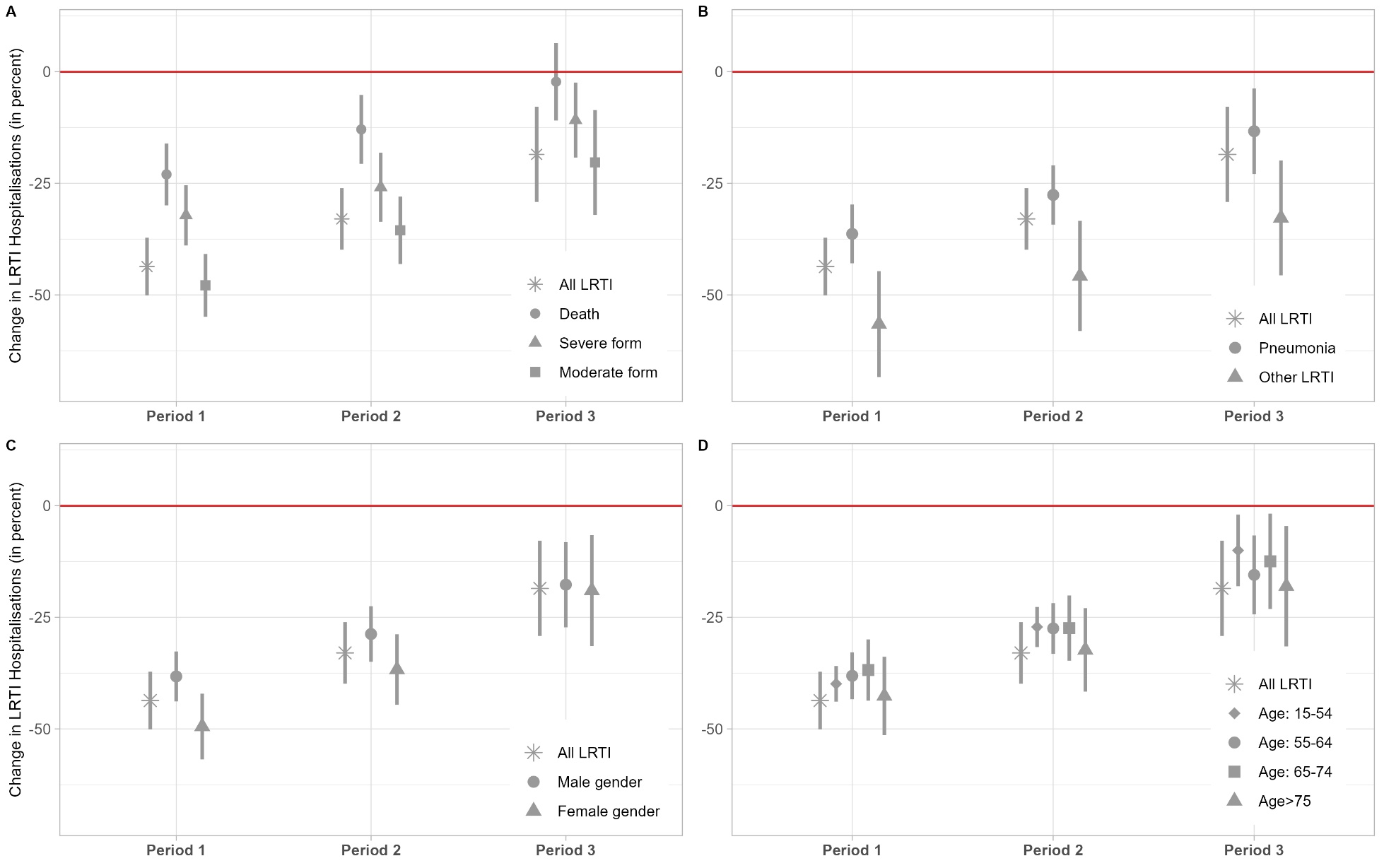

**Figure S3: Changes in LRTI hospitalisation rates during study periods according to the severity of the disease (A); type of LRTI (B); gender (C); age category (D)**

Period 1 extends from April 2020 to May 2021, period 2 from June 2021 to June 2022 and period 3 from July 2022 to June 2023.

Abbreviations: LRTI: Lower respiratory tract infection.

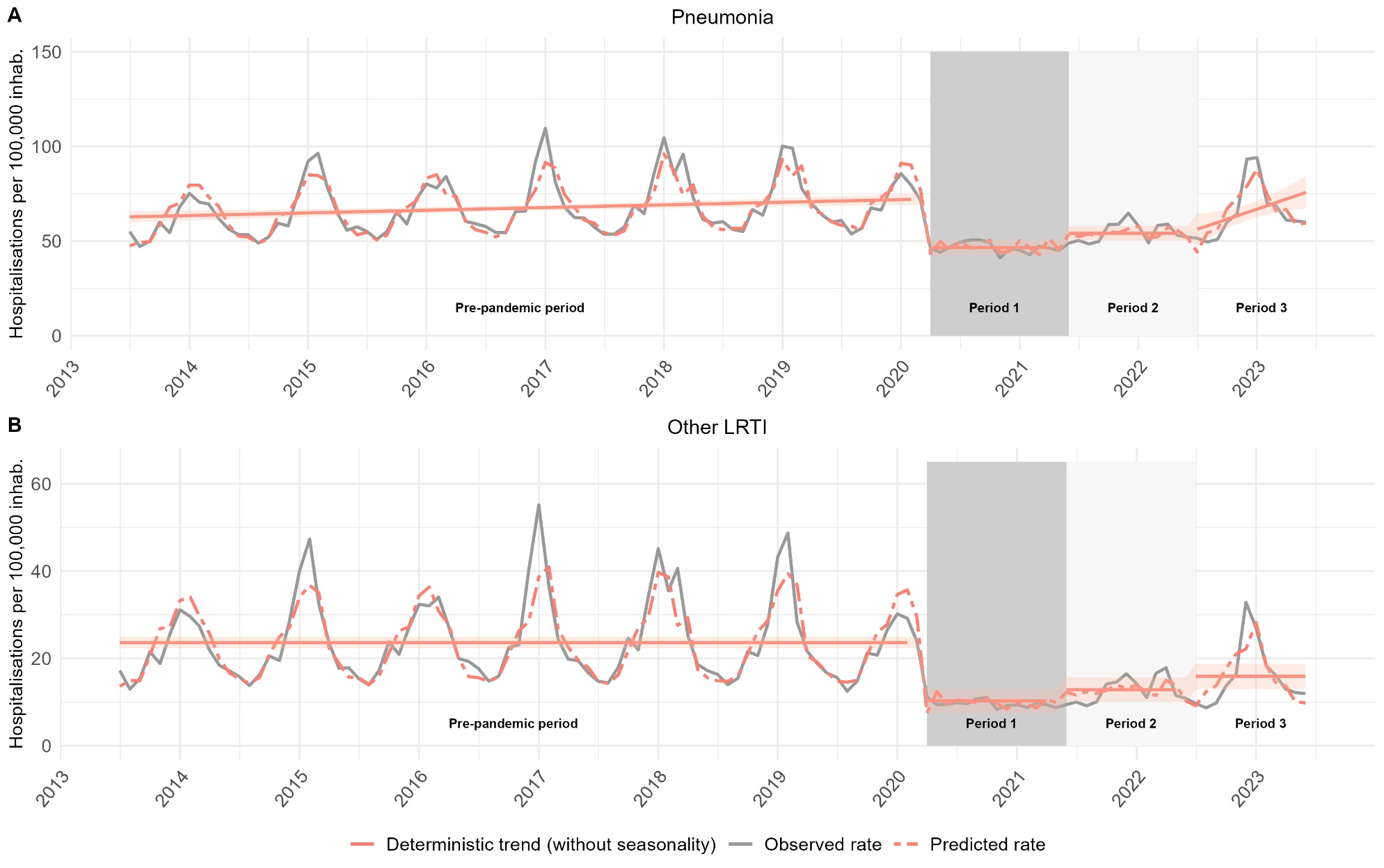

**Figure S4:** **Monthly hospitalisation rates of LRTI, contrasting pneumonia and other LRTI**

The observed incidence is depicted by the grey line. The model prediction is presented by the dotted red line, the predicted deterministic trend (regression line) and its 95% confidence intervals by the solid red line.

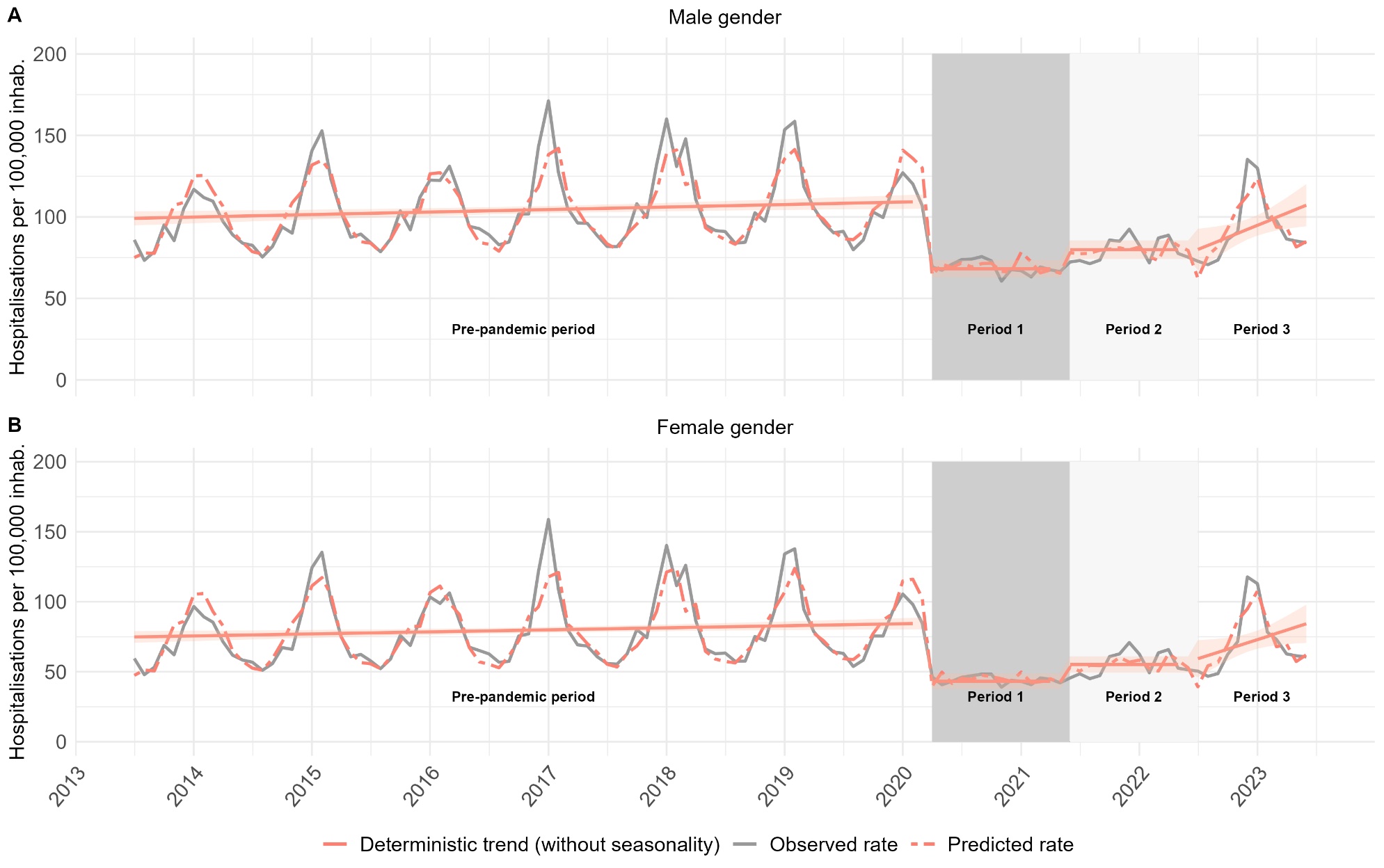

**Figure S5: Monthly hospitalisation rates of LRTI, by gender**

The observed incidence is depicted by the grey line. The model prediction is presented by the dotted red line, the predicted deterministic trend (regression line) and its 95% confidence intervals by the solid red line.

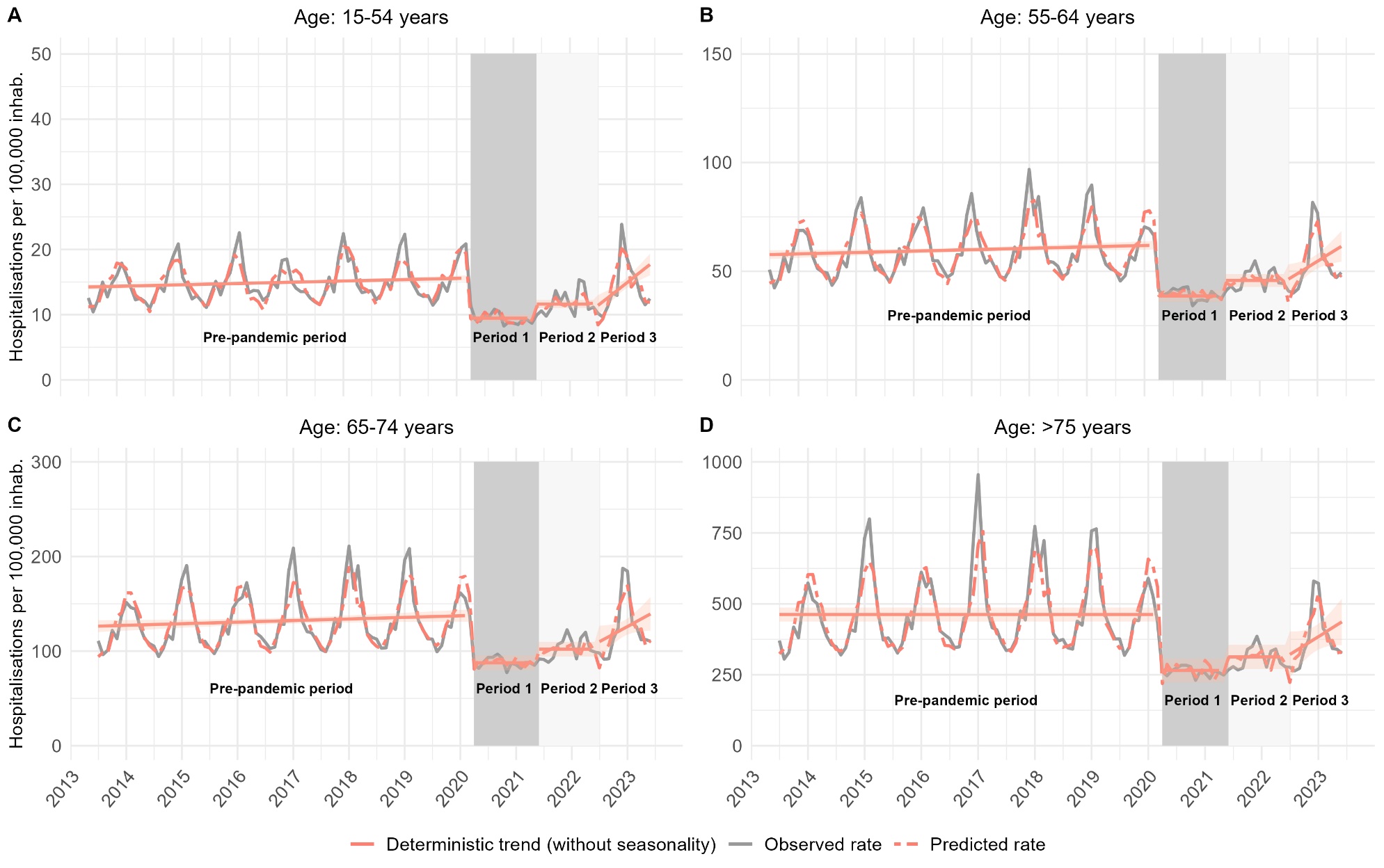

**Figure S6: Monthly hospitalisations rates of LRTI, by age category**

The observed incidence is depicted by the grey line. The model prediction is represented by the dotted red line, the predicted deterministic trend (regression line) and its 95% confidence intervals by the solid red line.

**Table S6**. **Estimations (Standard errors) and p value of the parameters included in the model for each series**

| **Parameters** | **All LRTI Model** | | **All LRTI**  **(with COVID-19 at period 3)** | | **Pneumonia** | | **Other LRTI** | | **CRD** | | **No CRD** | |
| --- | --- | --- | --- | --- | --- | --- | --- | --- | --- | --- | --- | --- |
| *Regression and seasonal parameters* | | | | | | | | | | | | |
| β_00_  β_10_  β_01_  β_11_  β_02_  β_12_  β_03_  β_13_  γ_0_  δ_0_  γ_3_  δ_3_ | 86.32 (2.12)  0.13 (0.05)  -41.60 (3.65)  -  -31.81 (3.91)  -  -32.47 (8.19)  2.25 (1.07)  -18.74 (1.72)  -19.37 (1.70)  -23.35 (4.83)  - | p<0.001  p=0.001  p<0.001  -  p<0.001  -  p<0.001  p=0.037  p<0.001  p<0.001  p<0.001  - | 86.25 (2.14)  0.13 (0.05)  -42.74 (3.92)  -  -33.21 (4.18)  -  0.19 (4.80)  -  -18.75 (1.74)  -19.38 (1.72)  -26.87 (4.88)  - | p<0.001  p=0.006  p<0.001  -  p<0.001  -  p=0.97  -  p<0.001  p<0.001  p<0.001  - | 62.06 (1.76)  0.12 (0.04)  -26.21 (2.53)  -  -20.10 (3.07)  -  -19.19 (5.32)  1.38 (0.64)  -11.27 (1.00)  -11.62 (0.99)  -15.42 (3.06)  - | p<0.001  p=0.001  p<0.001  -  p<0.001  -  p=0.001  p=0.03  p<0.001  p<0.001  p<0.001  - | 23.64 (0.67)  -  -13.36 (1.54)  -  -10.81 (1.59)  -  -7.74 (1.63)  -  -7.33 (1.07)  -7.35 (1.06)  -6.59 (2.22)  - | p<0.001  -  p<0.001  -  p<0.001  -  p<0.001  -  p<0.001  p<0.001  p=0.004  - | 31.21 (0.64)  -  -11.86 (1.19)  -  -7.71 (1.32)  -  -7.47 (2.56)  0.71 (0.33)  -6.11 (0.95)  -6.20 (0.95)  -6.92 (1.69)  - | p<0.001  -  p<0.001  -  p<0.001  -  p=0.004  p=0.04  p<0.001  p<0.001  p<0.001  - | 55.87 (1.49)  0.10 (0.03)  -28.19 (2.70)  -  -22.55 (2.90)  -  -21.85 (5.73)  1.26 (0.75)  -12.37 (1.22)  -12.79 (1.21)  -15.04 (3.41)  - | p<0.001  p=0.002  p<0.001  -  p<0.001  -  p<0.001  p=0.09  p<0.001  p<0.001  p<0.001  - |
| *Autoregressive parameters* | | | | | | | | | | | | |
| ɸ_1_  ɸ_2_  ɸ_10_  ɸ_11_  ɸ_12_  ɸ_13_  ɸ_14_  ɸ_18_ | -0.37 (0.09)  -0.31 (0.09)  -0.24 (0.09)  -  -  -  -  - | p<0.001  p=0.001  p=0.01  -  -  -  -  - | -0.32 (0.09)  0.29 (0.09)  0.26 (0.10)  -  -  -  -  - | p<0.001  p=0.001  p=0.008  -  -  -  -  - | -0.44 (0.09)  0.32 (0.09)  -  0.23 (0.09)  -0.40 (0.10)  0.34 (0.11)  -0.22 (0.10)  -0.20 (0.09) | p<0.001  p=0.001  -  p=0.02  p<0.001  p=0.002  p=0.03  p=0.03 | -0.47 (0.09)  0.29 (0.09)  -  -  -0.16 (0.09)  -  -  - | p<0.001  p=0.002  -  -  p=0.09  -  -  - | -0.37 (0.09)  0.18 (0.09)  -  -  -0.29 (0.10)  -  -  - | p<0.001  p=0.05  -  -  p=0.002  -  -  - | -0.40 (0.09)  0.33 (0.09)  0.23 (0.09)  -  -  -  -  - | p<0.001  p<0.001  p=0.01  -  -  -  -  - |
| *Residual variance* | | | | | | | | | | | | |
| σ^2^ | 105.84 | | 119.32 | | 37.49 | | 16.88 | | 10.16 | | 50.88 | |
| *Diagnostic tests (p-value)* | | | | | | | | | | | | |
| Independence**^1^**  Normality^2^ | 0.21  0.18 | | 0.38  0.21 | | 0.53  0.13 | | 0.75  0.11 | | 0.15  0.14 | | 0.28  0.13 | |
| **Parameters** | **COPD** | | **Asthma** | | **CF/Bronchiectasis** | | **ILD** | | **PVD** | | **Thoracic oncology** | |
| *Regression and seasonal parameters* | | | | | | | | | | | | |
| β_00_  β_10_  β_01_  β_11_  β_02_  β_12_  β_03_  β_13_  γ_0_  δ_0_  γ_3_  δ_3_ | 17.83 (0.36)  -  -7.45 (0.70)  -  -5.08 (0.76)  -  -5.23 (1.46)  0.40 (0.19)  -3.56 (0.53)  -3.70 (0.54)  -3.81 (0.96)  - | p<0.001  -  p<0.001  -  p<0.001  -  p=0.001  p=0.04  p<0.001  p<0.001  p<0.001  - | 3.02 (0.05)  -  -1.61 (0.11)  -  -0.91 (0.12)  -  -1.04 (0.31)  0.14 (0.04)  -0.88 (0.08)  -0.87 (0.08)  -1.41 (0.19)  - | p<0.001  -  p<0.001  -  p<0.001  -  p=0.001  p=0.001  p<0.001  p<0.001  p<0.001  - | 1.98 (0.10)  0.004 (0.001)  -0.89 (0.09)  -  -0.72 (0.13)  -  -0.83 (0.19)  0.05 (0.02)  -0.30 (0.04)  -0.34 (0.04)  -0.37 (0.09)  - | p<0.001  p=0.03  p<0.001  -  p<0.001  -  p<0.001  p=0.02  p<0.001  p<0.001  p<0.001  - | 1.85 (0.05)  -  -0.38 (0.07)  -  -0.36 (0.10)  -  -0.31 (0.18)  0.04 (0.02)  -0.33 (0.07)  -0.38 (0.07)  -0.39 (0.11)  - | p<0.001  -  p<0.001  -  p<0.001  -  p=0.08  p=0.08  p<0.001  p<0.001  p=0.001  - | 1.80 (0.11)  0.007 (0.002)  -0.79 (0.13)  -  -0.63 (0.17)  -  -0.43 (0.21)  -  -0.44 (0.09)  -0.37 (0.09)  -0.45 (0.13)  - | p<0.001  p=0.002  p<0.001  -  p=0.001  -  p=0.04  -  p<0.001  p<0.001  p=0.001  - | 3.50 (0.05)  0.008 (0.001)  -1.14 (0.10)  -  -0.95 (0.10)  -  -1.05 (0.17)  0.07 (0.02)  -0.32 (0.03)  -0.28 (0.03)  0.40 (0.09)  - | p<0.001  p<0.001  p<0.001  -  p<0.001  -  p<0.001  p=0.001  p<0.001  p<0.001  p<0.001  - |
| *Autoregressive parameters* | | | | | | | | | | | | |
| ɸ_1_  ɸ_2_  ɸ_10_  ɸ_11_  ɸ_12_  ɸ_13_  ɸ_15_  ɸ_24_ | -0.34 (0.09)  0.18 (0.09)  -  -  -0.30 (0.10)  -  -  - | p<0.001  p=0.06  -  -  p=0.003  -  -  - | -0.21 (0.08)  0.30 (0.08)  0.26 (0.09)  -  -  -  -  -0.29 (0.09) | p<0.001  p<0.001  p=0.004  -  -  -  -  p=0.002 | -0.22 (0.09)  -  -  -  -0.26 (0.10)  -  -0.28 (0.09)  - | p=0.01  -  -  -  p=0.007  -  p=0.003  - | -0.50 (0.09)  0.12 (0.09)  -  -  -0.27 (0.09)  -  -  - | p<0.001  p=0.19  -  -  p=0.004  -  -  - | -0.48 (0.09)  0.15 (0.09)  -  -  -0.34 (0.09)  -  -  - | p<0.001  p=0.10  p=0.001  -  -  - | -0.27 (0.09)  -  -  0.23 (0.09)  -0.30 (0.10)  0.26 (0.10)  -  - | p=0.004  -  -  p=0.02  p=0.003  p=0.009 |
| *Residual variance* | | | | | | | | | | | | |
| σ^2^ | 3.47 | | 0.18 | | 0.03 | | 0.03 | | 0.05 | | 0.03 | |
| *Diagnostic tests (p-value)* | | | | | | | | | | | | |
| Independence**^1^**  Normality^2^ | 0.22  0.15 | | 0.96  0.11 | | 0.51  0.20 | | 0.21  0.14 | | 0.29  0.18 | | 0.91  0.56 | |
| **Parameters** | **NMD with CRD** | | **Lung transplantation** | | **Death** | | **Severe form** | | **Moderate Form** | | **Male Gender** | |
| *Regression and seasonal parameters* | | | | | | | | | | | | |
| β_00_  β_10_  β_01_  β_11_  β_02_  β_12_  β_03_  β_13_  γ_0_  δ_0_  γ_3_  δ_3_ | 0.15 (0.002)  -  -0.06 (0.006)  -  -0.05 (0.006)  -  -0.03 (0.009)  -  -0.03 (0.003)  -0.02 (0.003)  -0.03 (0.009)  - | p<0.001  -  p<0.001  -  p<0.001  -  p<0.001  -  p<0.001  p<0.001  p=0.002  - | 0.18 (0.005)  .0001(.0001)  -0.07 (0.008)  -  -0.06 (0.009)  -  -0.05 (0.01)  -  -0.02 (0.003)  -0.02 (0.003)  -0.03 (0.009)  - | p<0.001  p<0.001  p<0.001  -  p<0.001  -  p<0.001  -  p<0.001  p<0.001  p=0.008  - | 11.71 (0.30)  -  -2.60 (0.44)  -  -1.50 (0.48)  -  -0.17 (0.53)  -  -1.72 (0.40)  -1.47 (0.39)  -1.55 (0.60)  - | p<0.001  -  p<0.001  -  p=0.002  -  0.75  -  p<0.001  p<0.001  p=0.01  - | 17.60 (0.38)  -  -5.66 (0.64)  -  -4.55 (0.73)  -  -1.90 (0.77)  -  -2.92 (0.56)  -2.62 (0.55)  -2.73 (0.87)  - | p<0.001  -  p<0.001  -  p<0.001  -  p=0.02  -  p<0.001  p<0.001  p=0.002  - | 57.50 (1.57)  0.09 (0.03)  -31.50 (2.86)  -  -23.89 (3.07)  -  -25.46 (6.13)  1.77 (0.80)  -13.79 (1.29)  -15.03 (1.27)  -17.85 (3.62)  - | p<0.001  p=0.006  p<0.001  -  p<0.001  -  p<0.001  p=0.03  p<0.001  p<0.001  p<0.001  - | 99.04 (2.22)  0.13 (0.05)  -42.08 (3.71)  -  -32.21 (4.14)  -  -35.21 (8.07)  2.34 (1.04)  -17.23 (1.91)  -18.31 (1.88)  -22.13 (4.64)  - | p<0.001  p=0.008  p<0.001  -  p<0.001  -  p<0.001  p=0.03  p<0.001  p<0.001  p<0.001  - |
| *Autoregressive parameters* | | | | | | | | | | | | |
| ɸ_1_  ɸ_2_  ɸ_10_  ɸ_12_  ɸ_24_ | -0.17 (0.09)  -  0.20 (0.10)  -  - | p=0.07  -  p=0.04  -  - | -  -  -  0.17 (0.10)  - | -  -  -  p=0.09  - | -0.48 (0.09)  0.15 (0.09)  -  -0.21 (0.09)  -0.17 (0.09) | p<0.001  p=0.08  -  p=0.02  p=0.06 | -0.54 (0.09)  0.21 (0.09)  -  -0.29 (0.09)  - | p<0.001  p=0.02  -  p=0.001  - | -0.35 (0.09)  0.31 (0.09)  0.24 (0.09)  -  - | p<0.001  p=0.001  p=0.009  -  - | -0.33 (0.09)  0.24 (0.09)  0.22 (0.09)  -  -0.18 (0.10) | p<0.001  p=0.008  p=0.02  -  p=0.07 |
| *Residual variance* | | | | | | | | | | | | |
| σ^2^ | 0.0003 | | 0.0005 | | 1.08 | | 2.15 | | 62.06 | | 97.99 | |
| *Diagnostic tests (p-value)* | | | | | | | | | | | | |
| Independence**^1^**  Normality^2^ | 0.58  0.62 | | 0.99  0.67 | | 0.47  0.20 | | 0.18  0.15 | | 0.32  0.12 | | 0.59  0.23 | |
| **Parameters** | **Female gender** | | **Age: 15-54 years** | | **Age: 55-64 years** | | **Age: 65-74 years** | | **Age: >75 years** | | **CRD (with COVID-19)** | |
| *Regression and seasonal parameters* | | | | | | | | | | | | |
| β_00_  β_10_  β_01_  β_11_  β_02_  β_12_  β_03_  β_13_  γ_0_  δ_0_  γ_3_  δ_3_ | 74.68 (2.14)  0.12 (0.05)  -42.27 (3.91)  -  -32.03 (4.18)  -  -30.78 (8.36)  2.13 (1.10)  -19.95 (1.78)  -20.26 (1.76)  -24.60 (4.98)  - | p<0.001  p=0.009  p<0.001  -  p<0.001  -  p<0.001  p=0.05  p<0.001  p<0.001  p<0.001  - | 14.23 (0.21)  0.02 (0.004)  -6.27 (0.38)  -  -4.36 (0.41)  -  -5.19 (0.98)  0.55 (0.13)  -2.17 (0.17)  -2.30 (0.17)  -4.77 (0.51)  - | p<0.001  p<0.001  p<0.001  -  p<0.001  -  p<0.001  p<0.001  p<0.001  p<0.001  p<0.001  - | 57.60 (1.05)  0.05 (0.02)  -23.74 (1.94)  -  -17.36 (2.08)  -  -18.38 (4.28)  1.30 (0.57)  -9.09 (0.85)  -10.58 (0.84)  -13.85 (2.44)  - | p<0.001  p=0.02  p<0.001  -  p<0.001  -  p<0.001  p=0.02  p<0.001  p<0.001  p<0.001  - | 126.25 (3.22)  0.14 (0.07)  -51.11 (5.68)  -  -38.60 (6.09)  -  -34.36(11.13)  2.54 (1.40)  -23.10 (2.66)  -24.65 (2.63)  -31.15 (6.38)  - | p<0.001  p=0.05  p<0.001  -  p<0.001  -  p=0.003  p=0.07  p<0.001  p<0.001  p<0.001  - | 462.40(12.5)  -  -197.09(21.9)  -  -149.29(23.3)  -  -151.21(47.5)  10.44 (6.3)  -101.45(19.1)  -101.27(19.0)  -97.78(32.4)  - | p<0.001  -  p<0.001  -  p<0.001  -  p=0.002  p=0.09  p<0.001  p<0.001  p<0.001  - | 31.14 (0.74)  -  -11.77 (1.26)  -  -7.64 (1.43)  -  1.9 (1.50)  -  -6.07 (1.07)  -6.10 (1.07)  -7.61 (1.75)  - | p<0.001  -  p<0.001  -  p<0.001  -  p=0.21  -  p<0.001  p<0.001  p<0.001  - |
| *Autoregressive parameters* | | | | | | | | | | | | |
| ɸ_1_  ɸ_2_  ɸ_10_  ɸ_11_  ɸ_12_  ɸ_24_ | -0.40 (0.09)  0.35 (0.09)  0.24 (0.09)  -  -  - | p<0.001  p<0.001  p=0.009  -  -  - | -  0.21 (0.08)  0.33 (0.09)  0.33 (0.09)  -  -0.22 (0.09) | -  p=0.008  p<0.001  p=0.001  -  p=0.02 | -0.30 (0.09)  0.27 (0.09)  0.31 (0.09)  -  -  - | p=0.001  p=0.003  p=0.001  -  -  - | -0.35 (0.09)  0.24 (0.09)  0.21 (0.10)  -  -0.16 (0.11)  - | p<0.001  p=0.01  p=0.04  -  p=0.13  - | -0.46 (0.09)  -0.24 (0.09)  -  -  -0.15 (0.09)  -0.19 (0.09) | p<0.001  p=0.01  -  -  p=0.10  p=0.04 | -0.37 (0.09)  0.16 (0.09)  -  -  -0.33 (0.10)  - | p<0.001  p=0.08  -  -  p=0.001  - |
| *Residual variance* | | | | | | | | | | | | |
| σ^2^ | 111.51 | | 1.48 | | 30.66 | | 173.50 | | 3475.79 | | 11.07 | |
| *Diagnostic tests (p-value)* | | | | | | | | | | | | |
| Independence^1^  Normality^2^ | 0.21  0.11 | | 0.27  0.71 | | 0.29  0.41 | | 0.65  0.55 | | 0.54  0.23 | | 0.27  0.65 | |
| **Parameters** | **No CRD**  **(with COVID-19)** | |  | |  | |  | |  | |  | |
| *Regression and seasonal parameters* | | | | | | | | | | | | |
| β_00_  β_10_  β_01_  β_11_  β_02_  β_12_  β_03_  β_13_  γ_0_  γ_0_  γ_3_  γ_3_ | 55.85 (1.51)  0.10 (0.03)  -28.30 (2.76)  -  -22.75 (2.94)  -  0.66 (3.37)  -  -12.38 (1.24)  -12.82 (1.22)  -17.75 (3.44)  - | p<0.001  p=0.002  p<0.001  -  p<0.001  -  p=0.85  -  p<0.001  p<0.001  p<0.001  - |  |  |  |  |  |  |  |  |  |  |
| *Autoregressive parameters* | | | | | | | | | | | | |
| ɸ_1_  ɸ_2_  ɸ_10_ | -0.33 (0.09)  0.31 (0.09)  0.25 (0.10) | p<0.001  p=0.001  p=0.01 |  |  |  |  |  |  |  |  |  |  |
| *Residual variance* | | | | | | | | | | | | |
| σ^2^ | 58.97 | |  |  |  |  |  |  |  |  |  |  |
| *Diagnostic tests (p-value)* | | | | | | | | | | | | |
| Independence**^1^**  Normality^2^ | 0.45  0.11 | |  | |  | |  | |  | |  | |

^1^Ljung-Box non correlation test up to lag 24; ^2^Shapiro-Wilk normality test (a maximum of 4% of residuals were removed to achieve normality).

Abbreviations: LRTI: Lower respiratory tract infection; CRD: chronic respiratory disease; ILD: interstitial lung disease; PVD: pulmonary vascular disease; NMD: neuromuscular disease.

**Table S7**. Proportion of COVID-19 infection among LRTI during and after the pandemic.

|  | **Period 1**,  April 2020 - May 2021  n= 831,627 | **Period 2**,  June 2021 - June 2022  n= 633,265 | **Period 3**,  July 2022 - June 2023  n= 651,763 |
| --- | --- | --- | --- |
| **COVID-19, n (%)** | 377,329 (45%) | 191,103 (30%) | 123,034 (19%) |
| **Other LRTI, n (%)** | 454,298 (55%) | 442,160 (70%) | 528,729 (81%) |

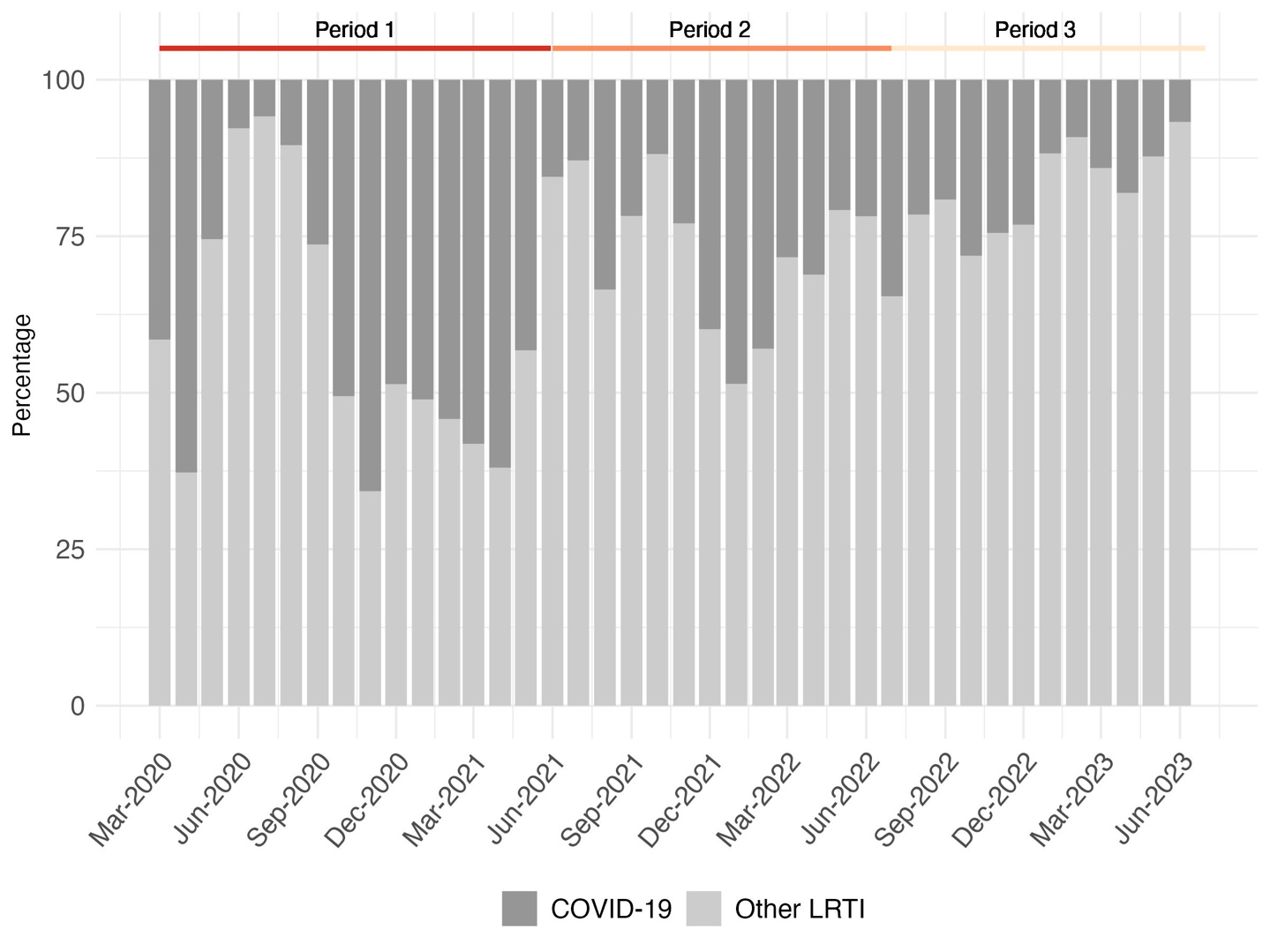

**Figure S7. Monthly distribution of the proportion of COVID-19 or other lower respiratory infections since the onset of the pandemic in France**

Period 1 extends from April 2020 to May 2021, period 2 from June 2021 to June 2022 and period 3 from July 2022 to June 2023

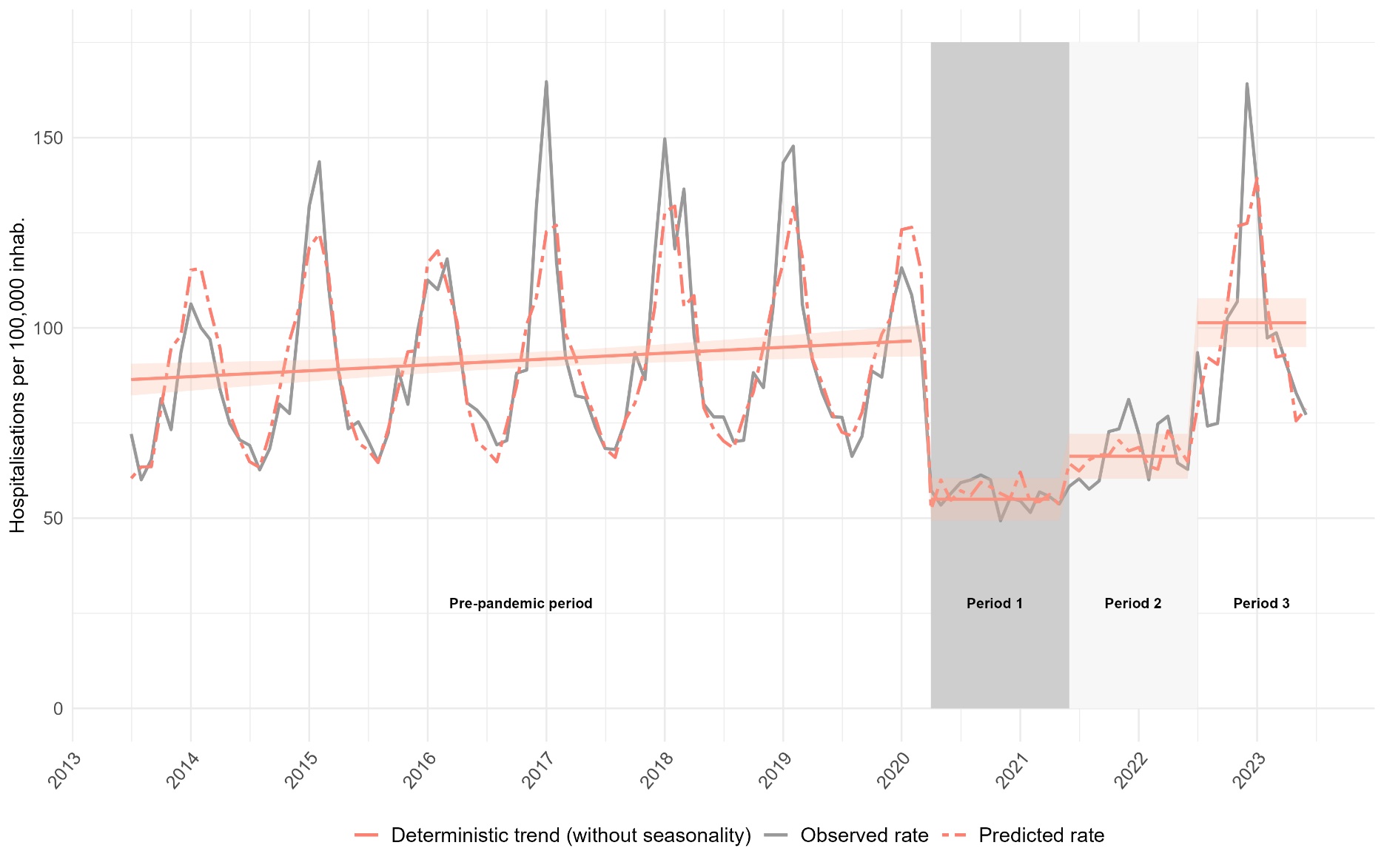

**Figure S8:** **Monthly hospitalisation rates of lower respiratory tract infection (including SARS-CoV-2 infection at period 3)**

The observed incidence is depicted by the grey line. The model prediction is represented by the dotted red line, the predicted deterministic trend (regression line) and its 95% confidence intervals by the solid red line.

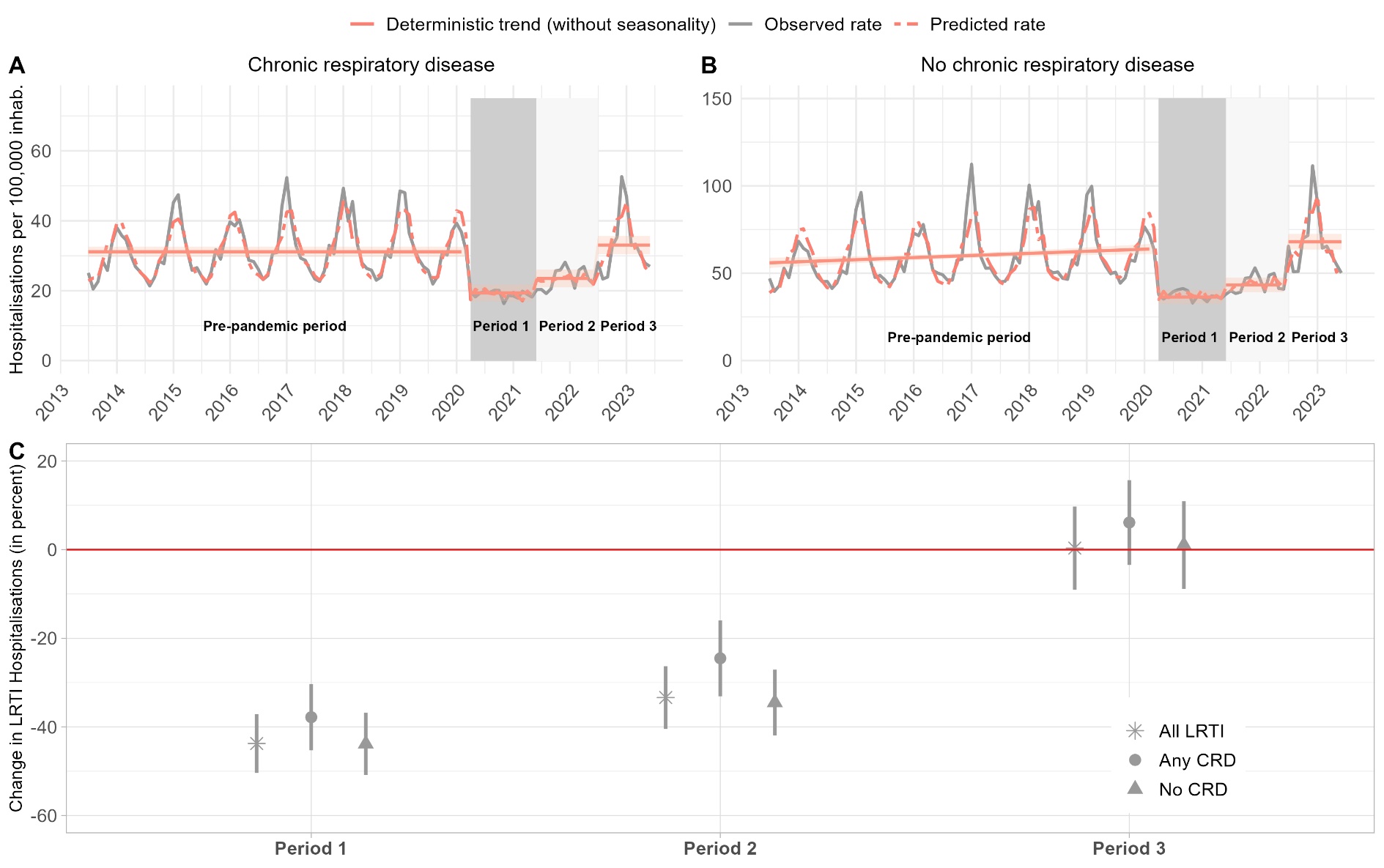

**Figure S9. Monthly hospitalisation rates of lower respiratory tract infection (A, B) and change in LRTI hospitalisations rates (C) relative to prior the pandemic period according to the presence of CRD (including SARS-CoV-2 infection at period 3)**

In figure A and B, the observed incidence is depicted by the grey line. The model prediction is represented by the dotted red line, the predicted deterministic trend (regression line) and its 95% confidence intervals by the solid red line. Period 1 extends from April 2020 to May 2021, period 2 from June 2021 to June 2022 and period 3 from July 2022 to June 2023.
